# Antimullerian Hormone Levels by Phenotypes in Polycystic Ovary Syndrome: A Systematic Review and Meta-Analysis

**DOI:** 10.1101/2025.01.25.25321110

**Authors:** Patrícia Jorge Schwenck de Carvalho, Giovana De Nardo Maffazioli, Ricardo Santos Simões, Renata Ferri Macchione, Sebastião Freitas de Medeiros, José Maria Soares, Edmund Chada Baracat, Gustavo Arantes Rosa Maciel

## Abstract

**Objective:** This study aims to evaluate antimüllerian hormone (AMH) levels across Polycystic Ovary Syndrome (PCOS) phenotypes (A, B, C, D) based on the Rotterdam diagnostic criteria.

**Data Sources:** A systematic review and meta-analysis were conducted following PRISMA guidelines. PubMed, Embase, ScienceDirect, and Web of Science were searched for studies published between January 2009 and June 2024.

**Study Eligibility Criteria:** Studies reporting AMH levels stratified by PCOS phenotypes based on the Rotterdam criteria, along with population characteristics and AMH measurement methods, were included. Animal studies, reviews, case reports, opinion articles, letters to the editor, other non-original research. and those lacking complete phenotype stratification were excluded.

**Study Appraisal and Synthesis Methods:** Study quality was assessed using the ROBINS-E tool. Primary outcomes included mean AMH levels across phenotypes, analyzed as tAMH (without assay differentiation) and bcAMH (Beckman Coulter Gen II assay). Secondary outcomes included age and BMI by phenotype. Random-effects meta-analysis was used to calculate mean levels and standardized mean differences (SMD) with 95% confidence intervals (CI). Heterogeneity was assessed using the I² statistic.

**Results:** Forty-nine studies involving 15,535 subjects revealed that AMH levels vary significantly by phenotype, being highest in A, followed by D, C, and B. For phenotype A: tAMH=9.87 ng/mL (SMD 2.97, CI 9.29–10.48); bcAMH=11.49 ng/mL (SMD 3.06, CI 10.47–12.61). For phenotype B: tAMH=5.71 ng/mL (SMD 0.98, CI 4.96–6.57); bcAMH=6.25 ng/mL (SMD 1.36, CI 5.20–7.51). There were no significant differences in age or BMI among phenotypes. High heterogeneity suggests additional factors influence AMH levels beyond phenotypic categories.

**Conclusions:** AMH levels are elevated in PCOS and vary across phenotypes, underscoring their potential to improve PCOS subtype characterization and guide management strategies. The findings highlight the need for standardizing AMH measurement techniques for consistent clinical application.

## Introduction

Polycystic ovary syndrome (PCOS) is a complex pathological condition characterized by hyperandrogenism, ovulatory dysfunction, and polycystic ovaries that affects women of reproductive age, with life-long impacts from adolescence to post-menopause [1]. It is a disorder, with an incidence varying from 4% to 10% of women in their reproductive years [2].

Different consensus of diagnostic criteria for PCOS were designed but the most widely accepted is the Rotterdam criteria (2003), which was updated by the latest international PCOS guidelines in 2023 [1]. According to the combination of Rotterdam criteria used for the diagnosis of PCOS, it can be classified into phenotypic presentations: phenotype A (PCOS-A) - hyperandrogenism (HA), oligo-amenorrhea, and polycystic ovarian morphology (PCOM) on ultrasound (US); phenotype B (PCOS-B) - HA and oligo-amenorrhea; phenotype C (PCOS-C) - HA and PCOM; phenotype D (PCOS-D) - oligo-amenorrhea and PCOM [3].

Although the Rotterdam diagnostic criteria are widely accepted, some points are still matters of controversy: the assessment of clinical and laboratory parameters of hyperandrogenism and hyperandrogenemia, the definition of oligo-amenorrhea throughout the reproductive age, the criteria to define the polycystic appearance of the ovaries on the ultrasound, the role of anti-müllerian hormone (AMH) in the diagnosis and phenotypes, among others, make its management, a clinical challenge [4]. There is a trend in the search for objective diagnostic criteria that encourages research into biomarkers in PCOS.

A promising diagnostic parameter is the anti-Müllerian hormone (AMH), a dimeric glycoprotein that plays an important role in sexual differentiation and regulation of folliculogenesis. It is named for its ability to inhibit the development of the Müllerian ducts in male fetuses[5]. AMH regulates ovarian folliculogenesis by inhibiting the recruitment of primordial follicles [6]. There are several possible uses for the AMH assessment: evaluation of the ovarian reserve, prediction of controlled ovarian stimulation, prediction of the natural age of menopause, assessment of the ovarian function, differentiation of some disorders of sex development, and tumor marker [7]. It is known that women with PCOS present elevated levels of AMH, correlating with testosterone levels, free androgen index, LH, average ovarian volume, the number of follicles on transvaginal ultrasound, and also with ovulatory dysfunction, including oligomenorrhea and amenorrhea [5]. As the levels of anti-Müllerian hormone can be influenced by each of the Rotterdam PCOS consensus diagnostic criteria, the various phenotypes present distinct AMH values, which may impact the use of this hormone as a diagnostic indicator for PCOS [8].

## Objectives

This systematic review aims to comprehensively evaluate and compare the mean levels of antimüllerian hormone (AMH) among the different phenotypes (A, B, C, and D) of Polycystic Ovary Syndrome (PCOS). Additionally, we aim to explore potential factors that may influence AMH levels within each phenotype, assess heterogeneity across studies, and conduct a meta-analysis to provide summary estimates of AMH levels in each PCOS phenotype.

## Methods

### a. Eligibility criteria, information sources, search strategy

The present research has been registered on the PROSPERO platform under the number CRD42023444193 and follows the Preferred Reporting Items for Systematic Reviews and Meta-analysis (PRISMA) statement [9]. This systematic review includes studies that meet the following criteria: women diagnosed with Polycystic Ovary Syndrome (PCOS) based on the Rotterdam criteria; studies that specifically report Anti-Mullerian Hormone (AMH) levels categorized into PCOS phenotypes A, B, C, and D; studies published in English, Spanish, Portuguese or French; both prospective and retrospective studies; and all types of study designs (randomized controlled trials, cohort studies, case-control studies, cross-sectional studies) as long as they provide the necessary data.

The exclusion criteria involve studies that do not include populations diagnosed with PCOS according to the Rotterdam criteria. These studies do not provide specific data on AMH levels, age, and body mass index (BMI), studies that do not distinguish between different phenotypes of PCOS according to the Rotterdam criteria, animal studies, reviews, case reports, opinion articles, letters to the editor, and other non-original research.

Two independent reviewers performed a comprehensive search of PubMed, Embase, Science Direct, and Web of Science databases for studies published up to July 2024. The following search terms were applied: “PCOS” or “Polycystic Ovary Syndrome,” “AMH” or “Anti-Mullerian Hormone” and “Phenotype” or “Phenotypic”. In addition, the reference lists of relevant publications were manually searched to include further relevant studies.

### b. Study selection

The reviewers screened independently the titles and abstracts of the retrieved articles based on the predefined inclusion and exclusion criteria. Full-text articles were obtained for potentially relevant studies. Any disagreements between the reviewers were resolved through discussion and consensus.

### c. Data extraction

Data extraction was performed independently by two reviewers using a standardized data extraction spreadsheet. Any discrepancies were resolved through discussion and consensus. Data collected included study characteristics (authors, publication year, study design, country of the study), participant characteristics - mean age and standard deviation (SD), mean BMI and SD by phenotype; mean AMH levels and SD by phenotype, and AMH assay used. In cases where specific age and BMI data for distinct phenotypes were not available, general age and BMI data from the PCOS control population were utilized. When articles reported only the median and interquartile range (iQR) without providing the standard deviation (SD), a formula was applied to estimate the mean and SD [11]. This approach allowed the inclusion of these studies in the meta-analysis, ensuring consistency in combining results across trials. In the studies where AMH levels were measured in pmol/L, we converted these values to ng/mL using the conversion factor, where 1 ng/mL is equivalent to 7.14 pmol/L [11]. To achieve greater reliability in the results, AMH values were converted to Beckman Coulter Generation II (Gen II) assay standards using validated formulas: Access AMH = 0.128 + 0.781 x Gen II for Access assay, Elecsys AMH = 0.253 + 0.688 x Ansh Labs and Elecsys AMH = 0.087 + 0.729 x Gen II for Ansh and Elecsys assays[11], Gen II = 1.40 x DSL - 0.62 for DSL [12], and Gen II = 1.353 × AMH (IOT) + 0.051 for Immunotech (IOT) [13]. The Elecsys formula was also applied to VIDAS AMH values due to their high correlation [14]. Studies that did not specify the AMH assay used or those without a conversion method were excluded from the final Gen II results. These conversion formulas were utilized for calculating both the mean AMH and standard deviation, ensuring consistency in measuring proportional variations across different assays.

### d. Assessment of risk of bias

The assessment of the risk of bias for the included studies was conducted using The Risk Of Bias In Non-randomized Studies - of Exposure (ROBINS-E) tool [15], a comprehensive instrument specifically tailored for evaluating bias in observational epidemiological studies, which evaluates seven domains including confounding, selection of participants, classification of interventions, deviations from intended interventions, missing data, measurement of outcomes, and selection of the reported result. Two independent reviewers applied the ROBINS-E tool to each study, with any discrepancies resolved through discussion or consultation with a third reviewer.

### e. Data synthesis

The statistical analysis was executed using the R software environment, incorporating the ’meta’, ’metaphor’, and ’ggplot2’ libraries for meta-analysis and data visualization. We opted for a random-effects model due to the expected study-to-study variability. The extent of heterogeneity was quantified using the I² statistic, which measures the percentage of variation across studies attributable to heterogeneity rather than chance. Additionally, we computed a 95% prediction interval to illustrate where the true effects of similar future studies are predicted to reside.

## Results

## a. Study selection

The literature search across all four databases yielded a total of 684 citations; of these, 201 were duplicates and were removed. The titles and abstracts of the remaining 483 citations were reviewed for relevance and screened against the inclusion and exclusion criteria. Ultimately, 82 full-text articles were assessed for eligibility. Of these, 35 were excluded, with the most common reason being the lack of specific data on AMH levels by the phenotypes of PCOS. In the end, 49 articles were included in this review, with 4 of them identified through citation searches. Further details on the search strategy and study selection process are presented in the PRISMA flow diagram (Figure 1).

**Figure 1.**
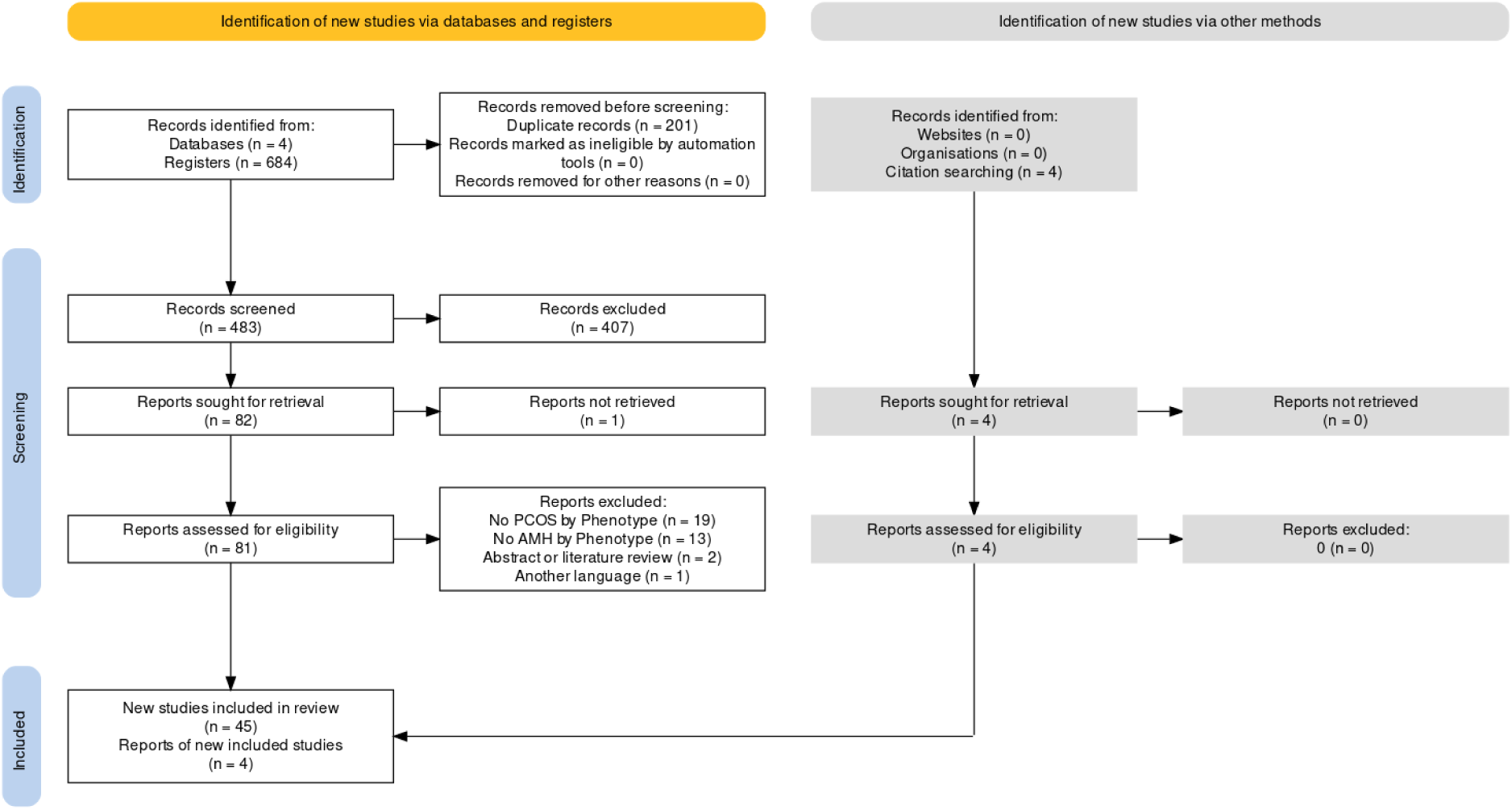
PRISMA flow diagram. Overview of study identification, screening, eligibility, and inclusion for the systematic review and meta-analysis.

### b. Study characteristics

The systematic review included a total of 49 studies, reflecting a variety of populations. The studies included were conducted between 2009 and 2024, with the number of PCOS women ranging from 31 to 3,775, amounting to a total of 15,388 subjects. The geographical distribution of these studies covered a comprehensive range of countries across all continents except Central and South America. The inclusion criteria for age varied among studies, with some specifying a range of 18-40 years, while others accepted a broader age group. Each study’s approach to the inclusion of infertile women also varied, with some studies explicitly including only non-infertile women, while others were inclusive or did not specify the fertility status of participants.

In terms of assay methods used to measure AMH levels, a variety of assays were employed. Several studies utilized the Beckman Coulter Generation II (GenII) assay, while others utilized the Roche Elecsys, ACCESS, Immunotech (IOT), DSL, and VIDAS. In some studies, the AMH assay was not determined. This variety in AMH assay methodologies show the importance of considering assay-specific differences when interpreting AMH levels across different populations.

Study characteristics are shown in Table 1.

**Table 1.** Characteristics of included studies. Summary of study year, country, sample sizes for total and PCOS phenotypes (A, B, C, D), AMH levels (ng/mL), and assay methods (BC: Beckman Coulter; ACCESS: Access Immunoassay; BC GEN II: Beckman Coulter Generation II; ELISA: Enzyme-Linked Immunosorbent Assay; ELECSYS: Roche Elecsys; IMMUNOTECH: Immunotech; DSL: Diagnostic Systems Laboratories; VIDAS: bioMérieux VIDAS; UNION: Union Immunoassay). [16], [17], [18], [19], [20], [21], [22], [23], [24], [25], [26], [27], [28], [29], [30], [31], [32], [33], [34], [35], [36], [37], [38], [39], [40], [41], [42], [43], [44], [45], [46], [47], [48], [49], [50], [51], [52], [53], [54], [55], [56], [57], [58], [59], [60], [61], [62], [63], [64].

| STUDY | YEAR | COUNTRY | N TOTAL PCOS | N PCOS A | N PCOS B | N PCOS C | N PCOS D | AMH ASSAY | AMH PCOS A ng/ml | AMH PCOS-A (ng/ml) | AMH PCOS-B (ng/ml) | AMH PCOS-C (ng/ml) | AMH PCOS-D (ng/ml) |
| --- | --- | --- | --- | --- | --- | --- | --- | --- | --- | --- | --- | --- | --- |
| Adamska 2020 | 2020 | Poland | 141 | 67 | 30 | 28 | 16 | BC | 9.6 | 9.96 | 9.33 | 9.1 | 9.03 |
| Albahlol 2023 | 2023 | Egypt | 185 | 88 | N/A | 41 | 56 | ACCESS | 9.44 | 9.44 | N/A | 8.58 | 11.16 |
| Alebić 2015 | 2015 | Croatia | 272 | 92 | N/A | 62 | 118 | BC GEN II | 7.51 | 7.42 | N/A | 4.69 | 6.68 |
| Amini 2018 | 2018 | Iran | 635 | 372 | 28 | 34 | 201 | N/A | 8.66 | 8.66 | 06.08 | 5.70 | 9.22 |
| Armstrong 2022 | 2022 | USA | 227 | 76 | 48 | 50 | 53 | ACCESS | 9.96 | 9.95 | 6.83 | 6.43 | 06.03 |
| Bahadur 2021 | 2021 | India | 307 | 114 | 25 | 18 | 150 | ELISA | 10.51 | 10.51 | 2.60 | 3.60 | 5.34 |
| Bell 2021 | 2021 | Australia | 31 | 7 | 10 | 12 | 2 | ELISA ANSH | 12.77 | 12.77 | 4.53 | 10.30 | 12.36 |
| Bhide 2017 | 2017 | UK | 192 | 91 | N/A | 42 | 59 | BC GEN II | 7.38 | 12.17 | N/A | 9.51 | 10.39 |
| Bozdag 2019 | 2019 | Turkey | 78 | 20 | 4 | 36 | 18 | ELECSYS | 5.64 | 5.64 | 2.20 | 3.20 | 3.10 |
| Cao NT 2018 | 2019 | Vietnam | 479 | 79 | 15 | 61 | 324 | ELECSYS | 9.81 | 9.81 | 9.11 | 8.65 | 8.74 |
| Carmina 2016 | 2016 | USA/Italy | 35 | N/A | N/A | 20 | 15 | BC GEN II | N/A | N/A | N/A | 5.5 | 5.4 |
| Carmina 2022 | 2022 | USA | 274 | 149 | 24 | 94 | 7 | BC GEN II | 10.2 | 10.2 | 5.2 | 6 | 5.3 |
| Cela 2018 | 2018 | Italy | 44 | 7 | 4 | 9 | 24 | N/A | 10.57 | 10.57 | 4.00 | 7.78 | 8.29 |
| De Vos 2018 | 2018 | Belgium | 295 | 66 | N/A | 54 | 175 | IMMUNOT ECH | 9.8 | 10.26 | N/A | 7.4 | 8.3 |
| Eftekhar 2019 | 2019 | Iran | 351 | 127 | 56 | 66 | 102 | N/A | 10.72 | 10.72 | 7.21 | 8.93 | 9.36 |
| Fraissinet 2017 | 2017 | France | 639 | 346 | 2 | 114 | 177 | BC GEN II | 10.08 | 10.08 | 3.59 | 7.43 | 7.72 |
| Gupta 2019 | 2019 | India | 57 | 57 | N/A | N/A | N/A | BC GEN II | 11.68 | 12.02 | N/A | N/A | N/A |
| Halder 2023 | 2023 | India | 131 | 87 | 23 | 8 | 13 | ELISA | 8.4 | 10.57 | 9.45 | 8.67 | 10.95 |
| Huijgen 2015 | 2015 | Netherlands | 218 | 94 | 9 | 9 | 106 | BC GEN II | N/A | N/A | N/A | N/A | 7.86 |
| Hwang 2013 | 2013 | Korea | 164 | 59 | N/A | N/A | 105 | IMMUNOT ECH | 10.1 | 10.1 | N/A | N/A | 10.0 |
| Ibrahim 2022 | 2022 | India | 200 | 50 | 50 | 50 | 50 | N/A | 9.43 | 9.43 | 7.13 | 8.53 | 9.71 |
| Jamil 2016 | 2016 | Iraq | 263 | 139 | 10 | 36 | 78 | BC GEN II | 5.62 | 5.62 | 03.01 | 5.53 | 5.25 |
| Le NSV 2021 | 2021 | Vietnam | 119 | 6 | 2 | 16 | 95 | ELECSYS | 7.35 | 7.35 | 2.73 | 7.94 | 7.35 |
| Mackens 2020 | 2020 | Belgium | 320 | 140 | N/A | 20 | 160 | ELECSYS | 12.4 | 12.4 | N/A | 7.7 | 10.4 |
| Mahajan 2019 | 2019 | India | 133 | 64 | 4 | 35 | 30 | ELECSYS | 09.05 | 09.05 | 3.32 | 6.31 | 6.39 |
| Malhotra 2023 | 2023 | India | 608 | 273 | 31 | 74 | 230 | ELISA<br>ANSH | 13.28 | 12.28 | 10.52 | 12.45 | 11.42 |
| Mitra 2023 | 2023 | India | 144 | 91 | 6 | 5 | 42 | N/A | 11.66 | 11.66 | 06.08 | 8.89 | 12.28 |
| Ozay 2020 | 2020 | Turkey | 350 | 117 | 89 | 72 | 72 | CUSABIO<br>ELISA | 9.17 | 9.17 | 8.15 | 7.30 | 6.18 |
| Piouka 2009 | 2009 | Greece | 100 | 25 | 25 | 25 | 25 | DSL | 9.2 | 9.2 | 6.3 | 5.1 | 5.3 |
| Rachmawati 2023 | 2023 | Indonesia | 43 | 17 | 12 | 7 | 7 | N/A | 10.31 | 10.31 | 4.18 | 07.02 | 7.51 |
| Ramezanali 2016 | 2016 | Iran | 386 | 168 | 103 | 83 | 32 | BC GEN II | 6.8 | 6.8 | 5.1 | 6.3 | 5.8 |
| Romualdi 2016 | 2016 | Italy | 117 | 73 | 10 | 13 | 21 | BC GEN II | 9.27 | 9.27 | 04.05 | 5.87 | 7.62 |
| Sachdeva 2019 | 2019 | India | 164 | 111 | 18 | 29 | 6 | ELECSYS | 11.34 | 11.34 | 8.69 | 9.44 | 7.76 |
| Sahmay 2013 | 2013 | Turkey | 251 | 119 | 26 | 45 | 61 | DSL | 9.50 | 9.50 | 03.06 | 6.12 | 8.2 |
| Sahmay 2018 | 2018 | Turkey | 286 | 204 | 16 | 51 | 15 | DSL | 7.64 | 8.34 | 2.43 | 6.61 | 9.28 |
| Santhiya 2021 | 2021 | India | 60 | 9 | 4 | 19 | 28 | VIDAS | 7.5 | 7.5 | 7.4 | 5.8 | 5.6 |
| Si M 2023 | 2023 | China | 1313 | 596 | 53 | 135 | 529 | N/A | 9.25 | 9.25 | 6.88 | 8.96 | 7.16 |
| Song 2017 | 2017 | Korea | 207 | 120 | 87 | N/A | N/A | BC | 18.6 | 17.7 | 9.7 | N/A | N/A |
| Sova 2019 | 2019 | Finland | 230 | 106 | N/A | N/A | 124 | VIDAS | 12.84 | 12.84 | N/A | N/A | 8.54 |
| Sumji 2023 | 2023 | India | 113 | 41 | 17 | 21 | 34 | ELISA<br>ANSH | 12.67 | 12.67 | 7.28 | 8.91 | 11.87 |
| Svetlana 2019 | 2019 | Russia | 100 | 53 | 27 | 15 | 5 | BC | 9.70 | 10.97 | 4.66 | 5.20 | 9.56 |
| Tian 2014 | 2014 | China | 160 | 40 | 40 | 40 | 40 | DSL | 8.33 | 8.33 | 4.29 | 5.49 | 6.70 |
| Urbanovych 2020 | 2020 | Ukraine | 80 | 18 | 19 | 20 | 23 | ELECSYS | 11.1 | 11.1 | 6.1 | 4.1 | 10.1 |
| Vural 2023 | 2023 | Turkey | 154 | 54 | 35 | 44 | 21 | ELECSYS | 7.8 | 7.8 | 5.7 | 5.8 | 5.5 |
| Wiweko 2014 | 2014 | Indonesia | 71 | 21 | 2 | 3 | 45 | BC GEN II | 11.1 | 11.1 | 11.5 | 8.72 | 8.2 |
| Wiweko 2018 | 2018 | Indonesia | 125 | 39 | 27 | 26 | 33 | BC GEN II | 11.7 | 13.33 | 11.33 | 11.96 | 6.6 |
| Yue CY 2018 | 2018 | China | 653 | 189 | 175 | 163 | 126 | UNION | 9.9 | 9.9 | 8.9 | 9.3 | 9.1 |
| Zhang 2023 | 2023 | China | 3775 | 2430 | 183 | N/A | 1162 | ELECSYS | 9.23 | 9.23 | 4.43 | N/A | 6.99 |
| Zhao 2023 | 2023 | China | 220 | 84 | 34 | N/A | 102 | N/A | 8.89 | 8.89 | 9.81 | N/A | 6.60 |

### c. Risk of bias of included studies

The risk of bias within the included studies was assessed using the ROBINS-E tool. The evaluation revealed that most studies maintained a low risk of bias across several domains, particularly in the measurement of exposure and outcome and reporting of results, reflecting methodological robustness. However, concerns were noted in certain areas. Bias due to participant selection and due to confounding were evident, with several studies showing some concerns and high risk, pointing to potential limitations in study design or execution. The overall risk of bias was moderated in the majority of studies, although some exhibited a high risk. This evaluation underscores the need for a cautious interpretation of the results.

The detailed findings of this risk assessment are presented in Figure 2.

**Figure 2.**
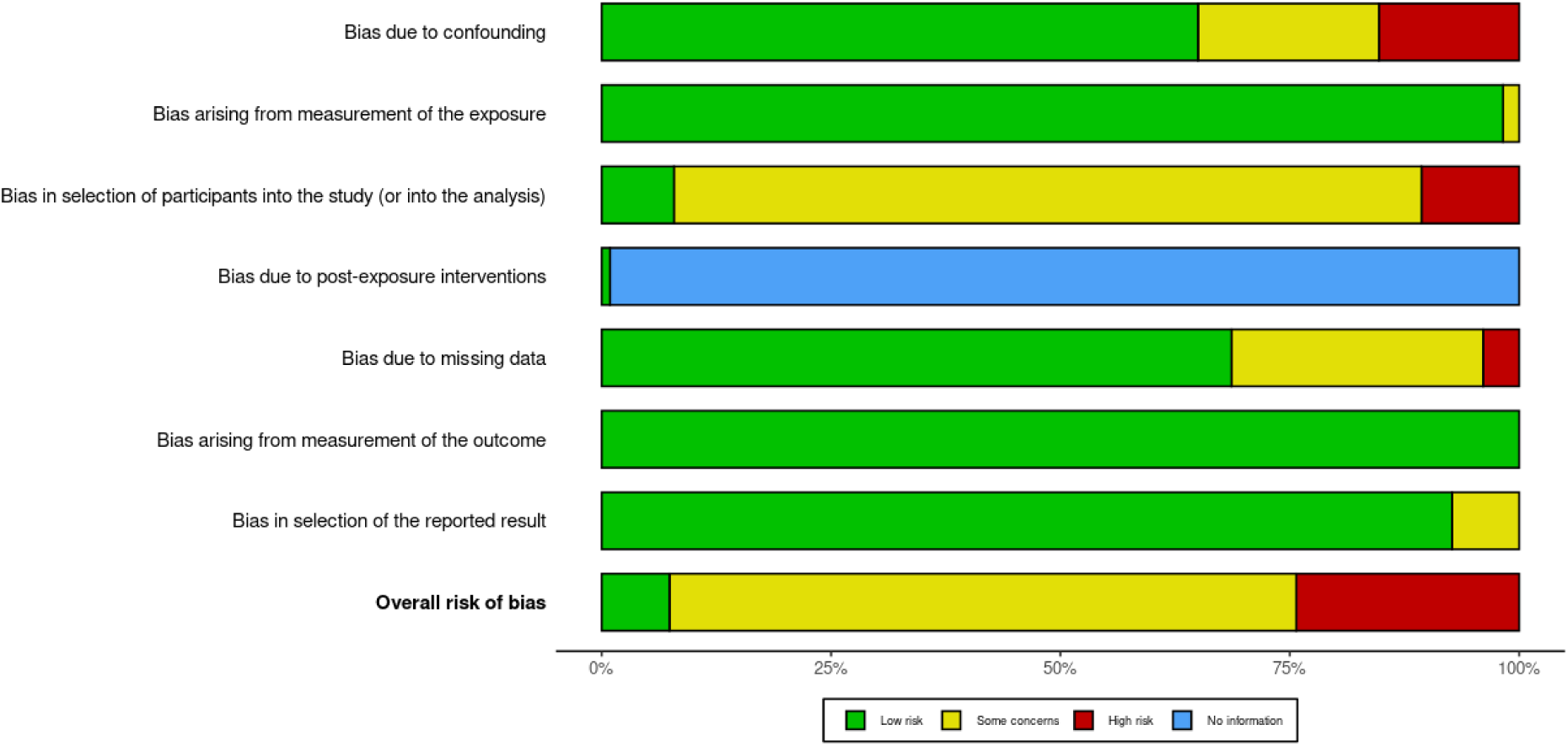
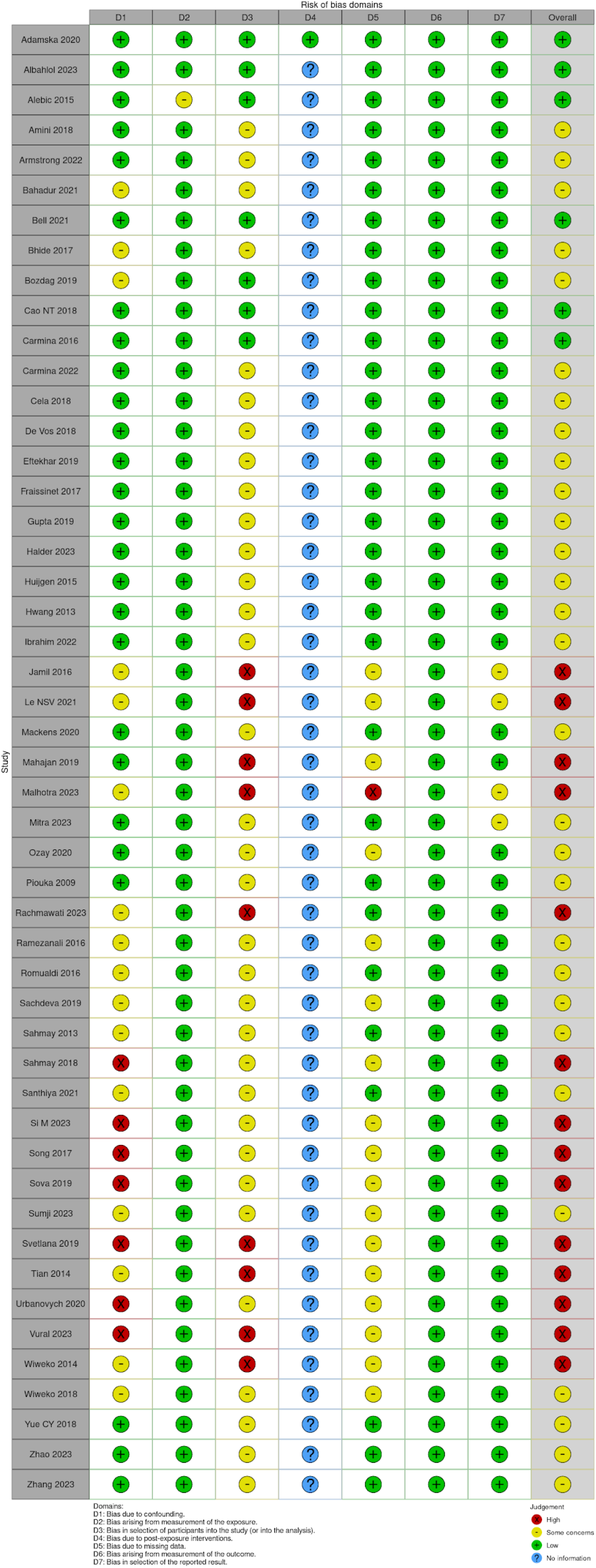
Risk of bias assessment using ROBINS-E. Combined graphical and tabular representation of the risk of bias evaluation for included studies. Figure 2A shows the proportion of studies categorized as low risk, some concerns, high risk, and no information across the specified domains. Figure 2B provides a detailed tabular overview of the risk of bias assessment for each study across all domains.

### d. Synthesis of results

The results from our systematic review highlight significant variability in AMH levels when stratified by PCOS phenotypes (figure 3). The **PCOS phenotype A** had the highest mean AMH level measured without assay differentiation at **9.87 ng/mL (SMD 2.97, CI 95% 9.29-10.48)**, and with the Beckman Coulter Gen II assay conversion at **11.49 ng/mL (SMD 3.06, CI 95% 10.47-12.61).** In contrast, **phenotype B** showed the lowest median AMH levels in both measurements, **5.71 ng/mL (SMD 0.98, CI 95% 4.96-6.57)** without assay differentiation and **6.25 ng/mL (SMD 1.36, CI 95% 5.20-7.51)** with Beckman Coulter Gen II. **PCOS C** mean AMH without assay differentiation **6.91 ng/mL (SMD 1.42, CI 95% 6.30-7.57)** and converted to **BC GENII 7.98 ng/mL (SMD 1.52, CI 95% 7.16-8.90)**, **PCOS D** without assay differentiation **7.80 ng/mL (SMD 2.27, CI 95% 7.20-8.45)** and **BC GEN II** 8.97 ng/mL (SMD 2.44, CI 95% 8.04-10.00).

**Figure 3.**
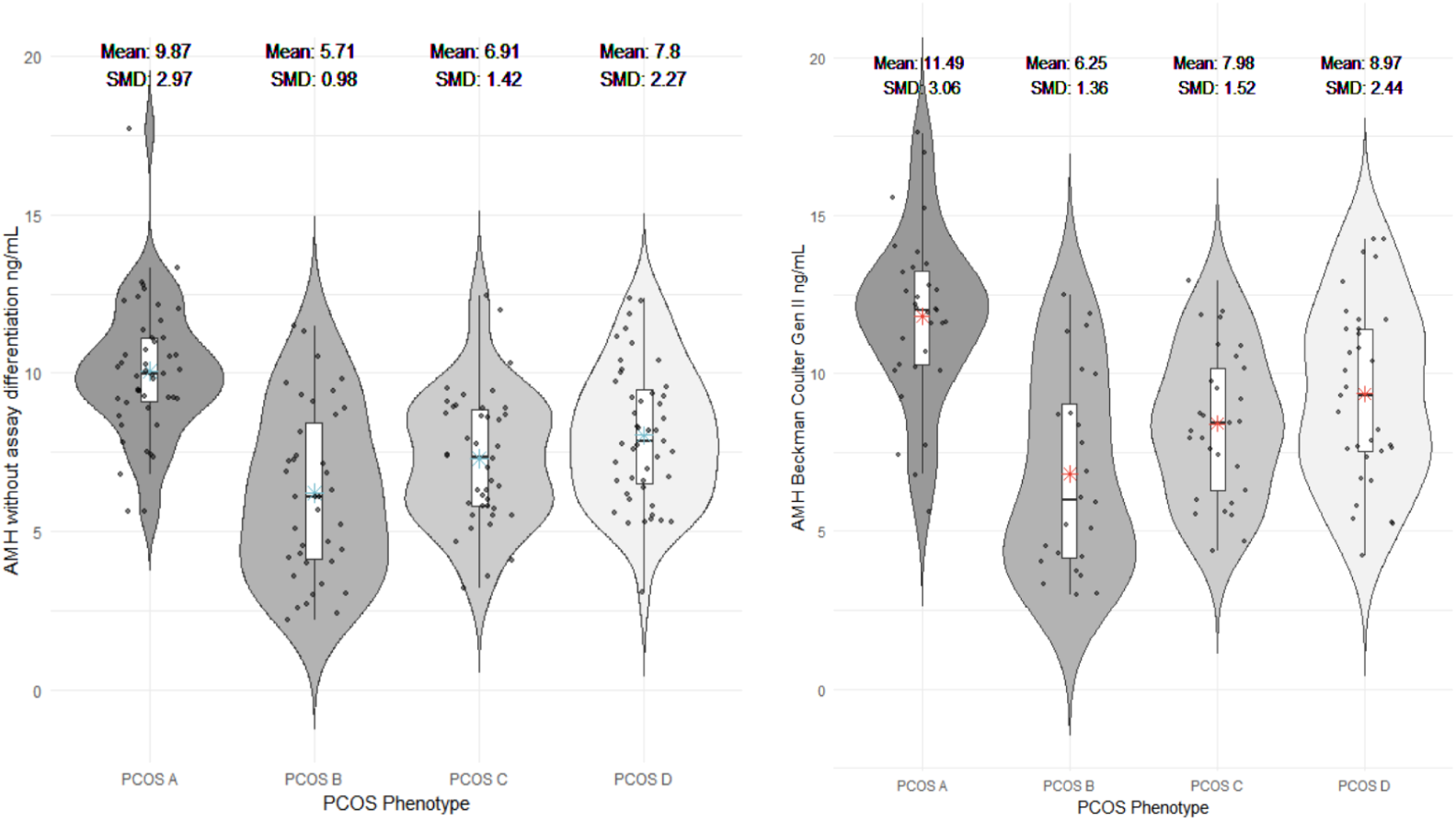
AMH levels across PCOS phenotypes. Violin plots displaying the distribution of AMH levels (ng/mL) across PCOS phenotypes A, B, C, and D, without assay differentiation (left) and after Beckman Coulter Gen II assay normalization (right). Mean values and standardized mean differences (SMD) are highlighted for each phenotype.

The age distribution across phenotypes was relatively similar, with means ranging from 26.17 to 27.57 years, indicating that age may not be a distinguishing factor in AMH levels among the included studies. The body mass index (BMI) also presented low differences, with the mean BMI being the lowest in phenotype D at 24.78 kg/m² and high in PCOS A (mean 26.11 kg/m²) (figure 4).

**Figure 4.**
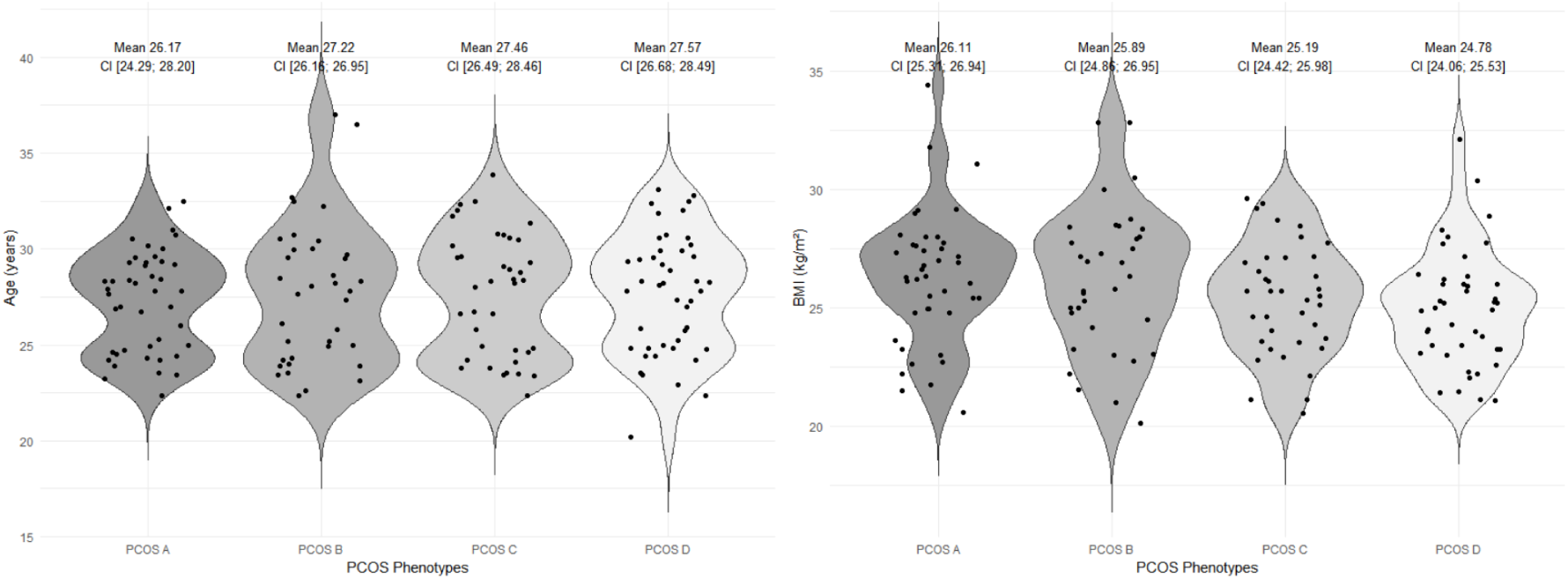
Age and BMI distribution across PCOS phenotypes. Violin plots illustrate the distribution of age (years) and body mass index (BMI, kg/m²) among PCOS phenotypes A, B, C, and D. Mean values with confidence intervals (CI) are displayed for each phenotype.

Our results showed notable disparities in sample sizes across the four PCOS phenotypes, reflecting the prevalence of each phenotype in the studied populations. Phenotype A displayed the largest sample sizes, suggesting that this phenotype is more commonly studied and also more prevalent. Larger sample sizes generally provide more precise estimates, as indicated by the narrower confidence intervals in our forest plots. This implies that our findings for Phenotype A might be the most reliable indicator of AMH levels among the PCOS phenotypes we studied.

In contrast, Phenotype B often had smaller sample sizes, indicating that this phenotype might be less common, less frequently studied, or more challenging to diagnose. As a result, while the estimated AMH levels for Phenotype B are valuable, they are subject to more variability and might require cautious interpretation and verification in larger cohorts. Phenotypes C and D had moderate sample sizes, providing a balance between the extremes of A and B.

Our meta-analysis has revealed a pronounced degree of heterogeneity in AMH levels, as evidenced by I² values close to or at 98% across all phenotypes, which is indicative of significant variability among the included studies. This heterogeneity is substantial and merits careful consideration. Small-study effects, methodological differences, or even publication bias could be influencing the results. Variations in study populations, ethnicity, AMH assays, and differences in PCOS symptoms might lead to variability in AMH levels. The significant heterogeneity underscores the complexity of PCOS as a syndrome with a spectrum of manifestations and the challenges in using AMH as a uniform biomarker.

Additional forest plots displaying subgroup analyses and details of the meta-analysis are provided in the supplementary material (Supplementary File 1).

**Figure 5.**
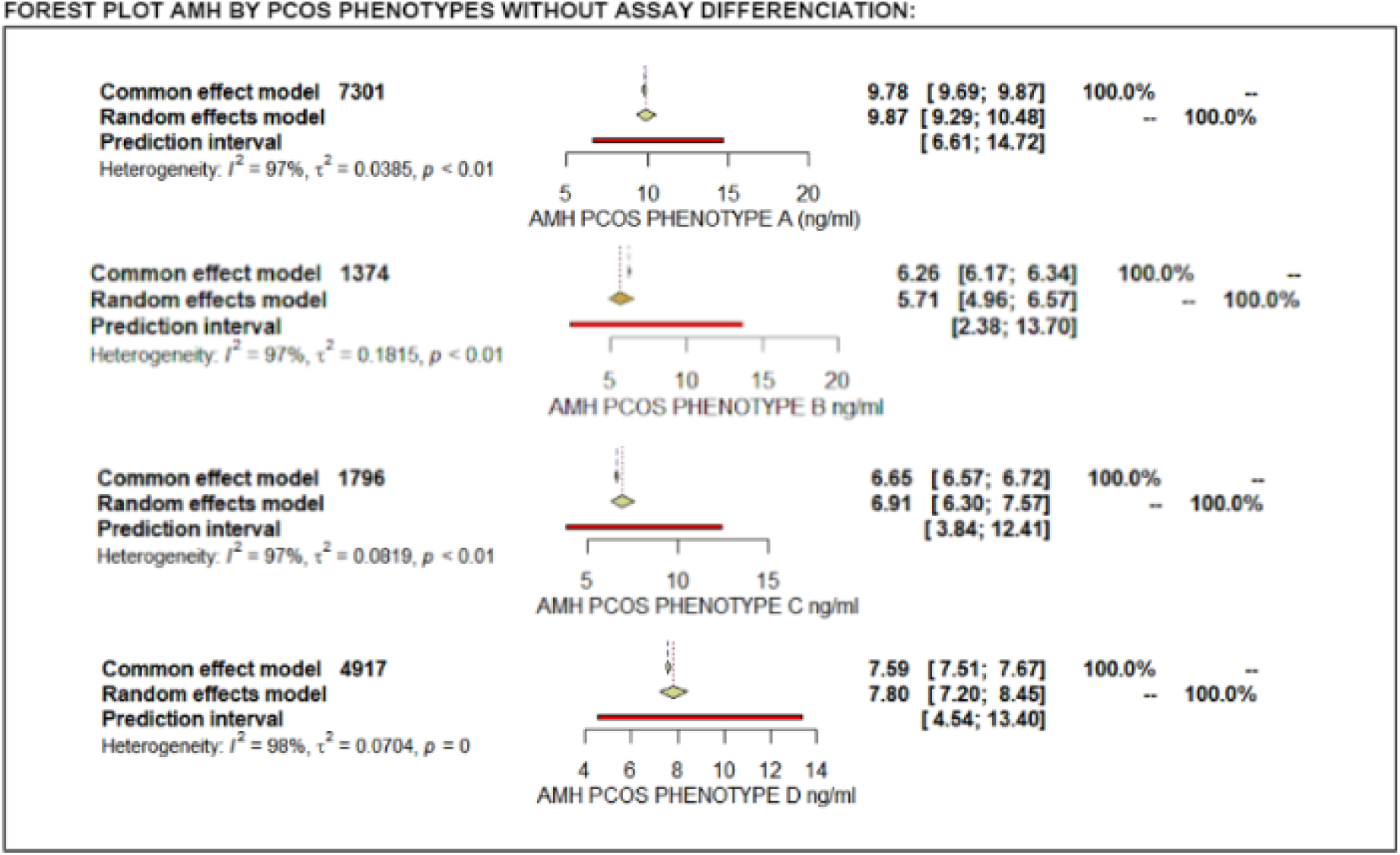
Forest plot of AMH levels by PCOS phenotypes without assay differentiation. Forest plot illustrating mean AMH levels (ng/mL) and 95% confidence intervals (CI) for PCOS phenotypes A, B, C, and D. Results were calculated using random-effects models.

**Figure 6.**
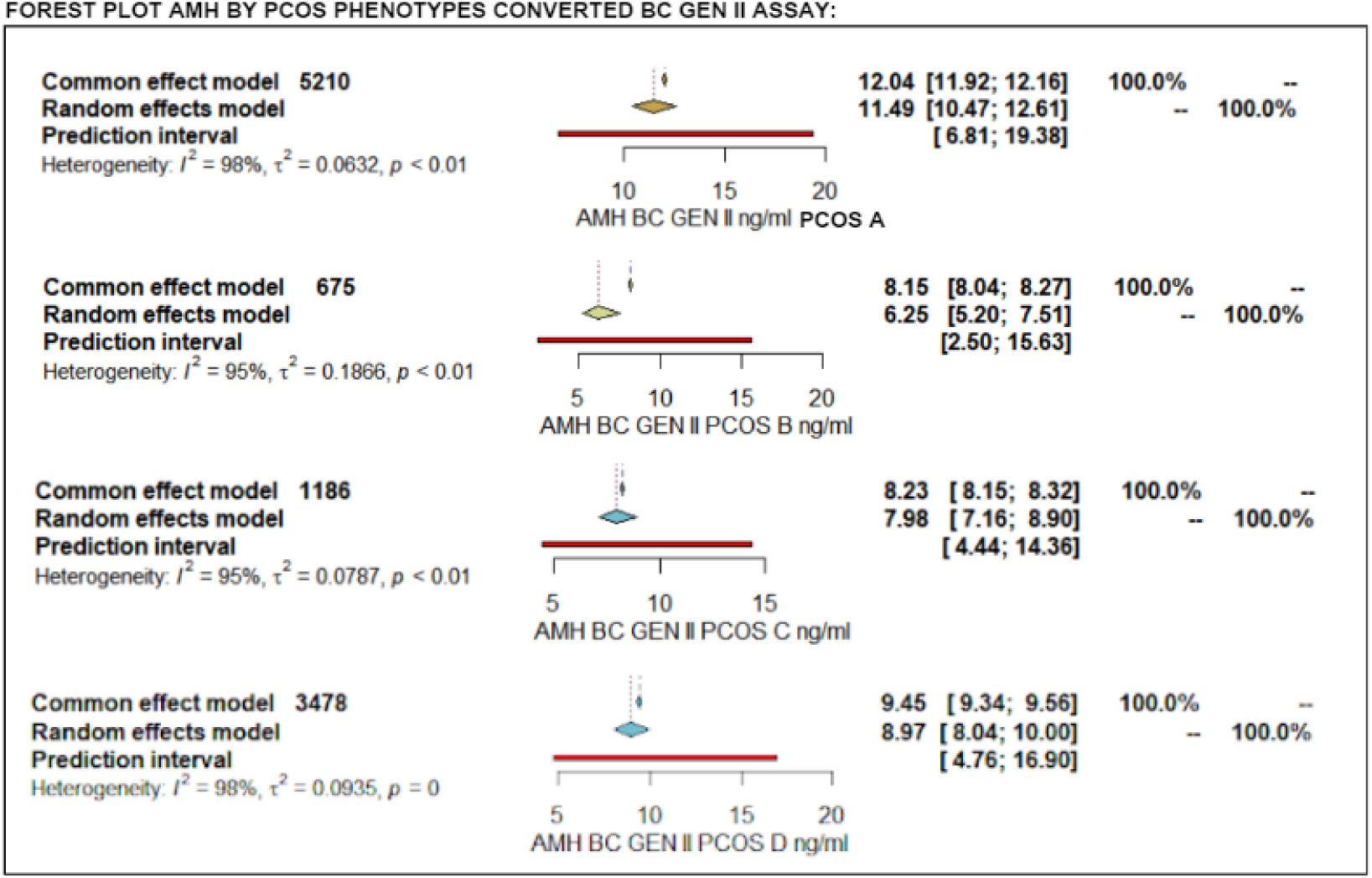
Forest plot of AMH levels by PCOS phenotypes converted to Beckman Coulter Gen II assay. Forest plot showing mean AMH levels (ng/mL) and 95% confidence intervals (CI) for PCOS phenotypes A, B, C, and D, standardized to Beckman Coulter Gen II assay values.

## Comment

### a. Principal Findings

Our meta-analysis found significant variability in anti-Müllerian hormone (AMH) levels across different phenotypes of polycystic ovary syndrome (PCOS). Phenotype A consistently exhibited the highest AMH levels, followed by phenotypes D, C, and B. These results align with previous research indicating that women with more severe ovulatory dysfunction or hyperandrogenism, particularly in phenotype A, tend to have elevated AMH levels [47];[16]. Interestingly, we observed that phenotype D had higher AMH levels than phenotype C, suggesting that oligoanovulation may exert a greater influence on AMH levels than hyperandrogenism alone. This is supported by studies showing that ovulatory dysfunction is closely linked to follicular excess, which in turn elevates AMH levels [24]. Conversely, phenotype B showed the lowest AMH levels, this supports the hypothesis that polycystic ovarian morphology is the primary factor driving elevated AMH levels in PCOS, as previously suggested by Carmina et. al. [26], who found that PCOM correlates more strongly with AMH than hyperandrogenism or oligoanovulation. This gradient in AMH levels suggests that AMH may serve as a useful clinical indicator for distinguishing between PCOS phenotypes, which could have implications for individualized treatment approaches.

### b. Comparison with Existing Literature

Previous meta-analyses, such as those by Anand et al. [65] and Tsukui et al. [66], have focused on the diagnostic accuracy of anti-Müllerian hormone (AMH) in distinguishing PCOS patients from controls, consistently showing elevated AMH levels in women with PCOS. A recent study showed the potential AMH as a diagnostic biomarker for polycystic ovary syndrome (PCOS). A serum AMH concentration of at least 5.39 ng/mL was associated with an increased risk of PCOS, with a sensitivity of 88.6%, specificity of 92.75%, an area under the curve (AUC) of 0.95. [67] However, these analyses did not differentiate between the various PCOS phenotypes, which we believe is crucial given the metabolic heterogeneity within the syndrome. As highlighted by Soares-Jr et al. (2023) [68], different PCOS phenotypes exhibit distinct metabolic profiles, with phenotype A, in particular, being associated with a higher risk of metabolic syndrome and type 2 diabetes. This reinforces the importance of stratifying PCOS by phenotypes to better understand the variability in clinical and metabolic outcomes. Our study adds to the literature by providing insights into the variability of AMH levels across these phenotypes, suggesting that polycystic ovarian morphology (PCOM) plays a more significant role in elevating AMH levels than other features, such as hyperandrogenism or ovulatory dysfunction.

### c. Strengths and Limitations

A major strength of this meta-analysis is the inclusion of a large sample size (15,535 women) across diverse populations, which enhances the generalizability of the findings. Additionally, the use of conversion formulas for standardizing AMH measurements across different assays adds reliability to the results. However, significant heterogeneity across studies, as indicated by high I² values, suggests that factors beyond phenotypic classification, such as ethnicity, assay methods, and inclusion criteria, may influence AMH levels. The variability in sample sizes across phenotypes, particularly the smaller cohort for phenotype B, represents another limitation that could affect the precision of our estimates for this group.

### d. Conclusions and Implications

Our findings suggest that AMH levels could serve as an important diagnostic indicator for differentiating between PCOS phenotypes, particularly when integrated with other clinical parameters [27]. The distinct AMH profiles across phenotypes underscore the need for phenotype-specific approaches in PCOS management. However, the variability in AMH assays across studies highlights the importance of standardized measurement techniques to ensure consistent clinical application [22]. Moving forward, future research should focus on reducing heterogeneity by standardizing AMH measurements and exploring the role of AMH in guiding personalized treatments for PCOS.

## Data Availability

All data produced in the present study are available upon reasonable request to the authors

