## Supplementary File 1 for "Antimullerian Hormone Levels by Phenotypes in Polycystic Ovary Syndrome: A Systematic Review and Meta-Analysis"

Detailed forest plots categorized by PCOS phenotype (A, B, C, D), showing age, BMI, AMH levels without assay differentiation, and AMH levels standardized to the Beckman Coulter assay.

### PCOS-A Age:

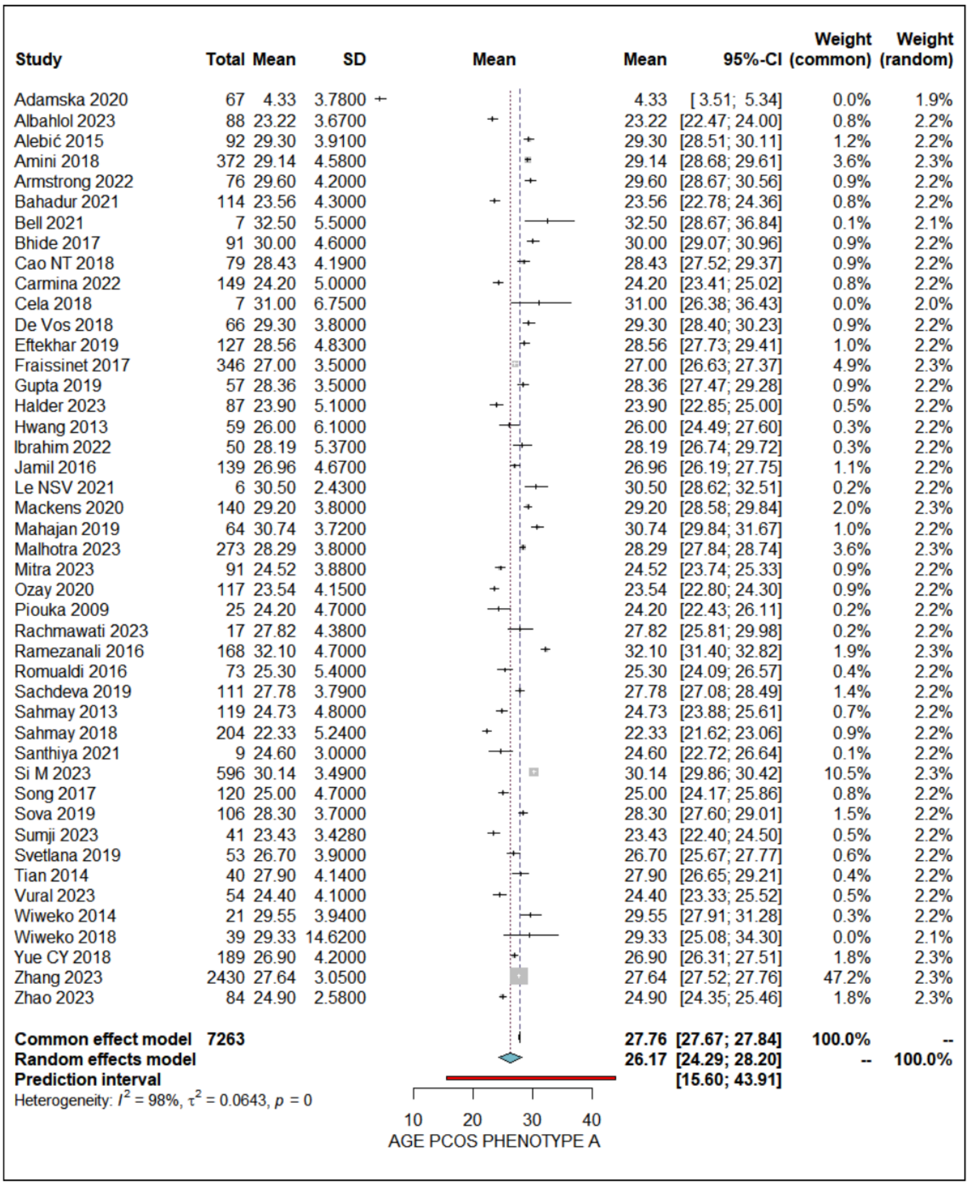


### PCOS-A BMI:

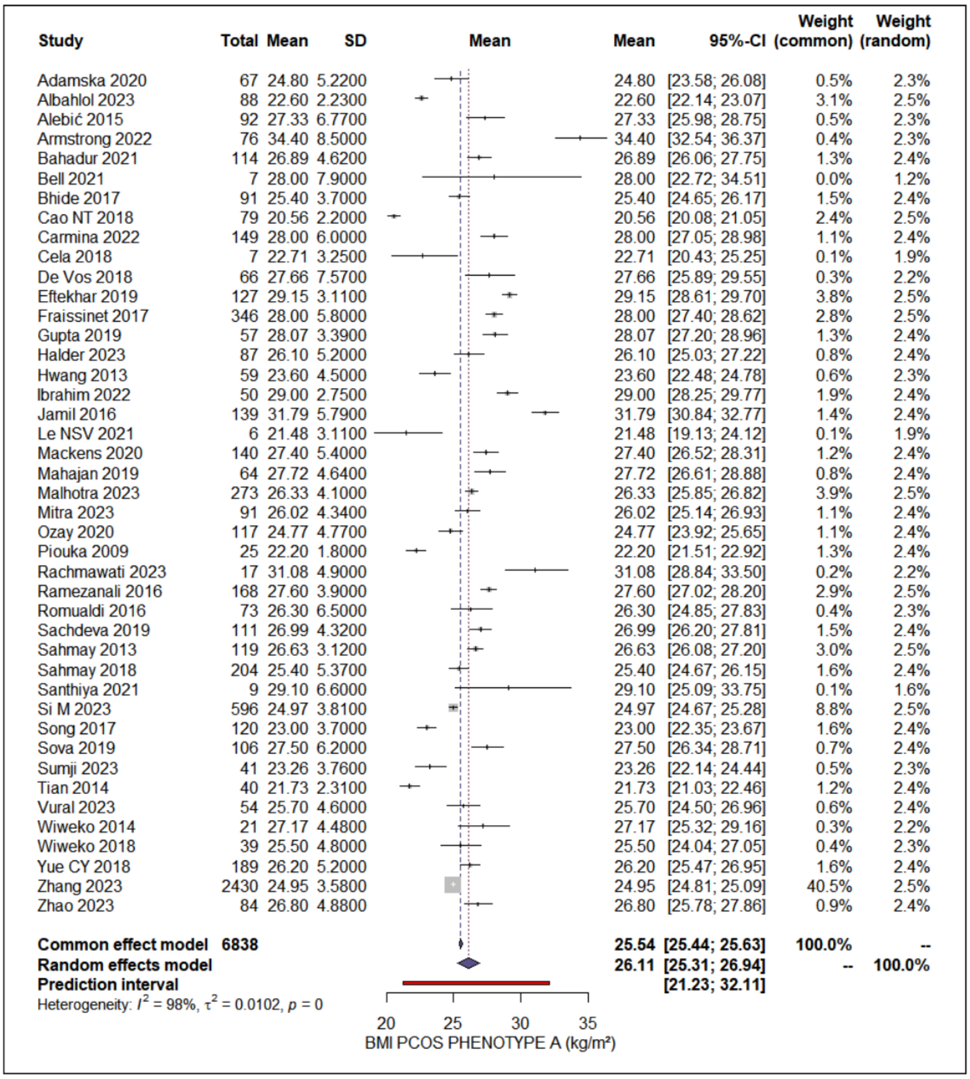


**PCOS-A AMH levels without assay differentiation:**

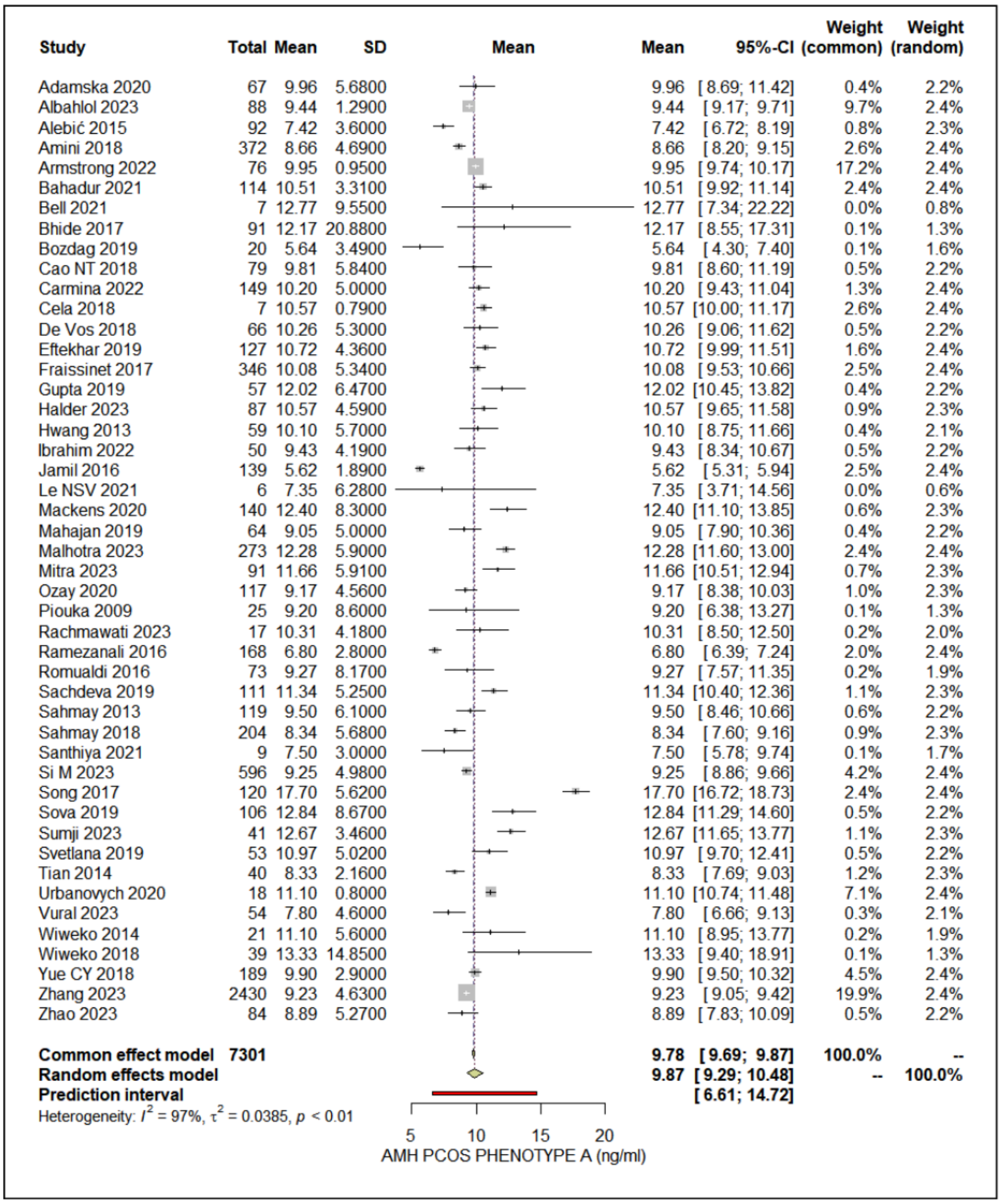


**PCOS-A AMH levels standardized to the Beckman Coulter Gen II assay:**

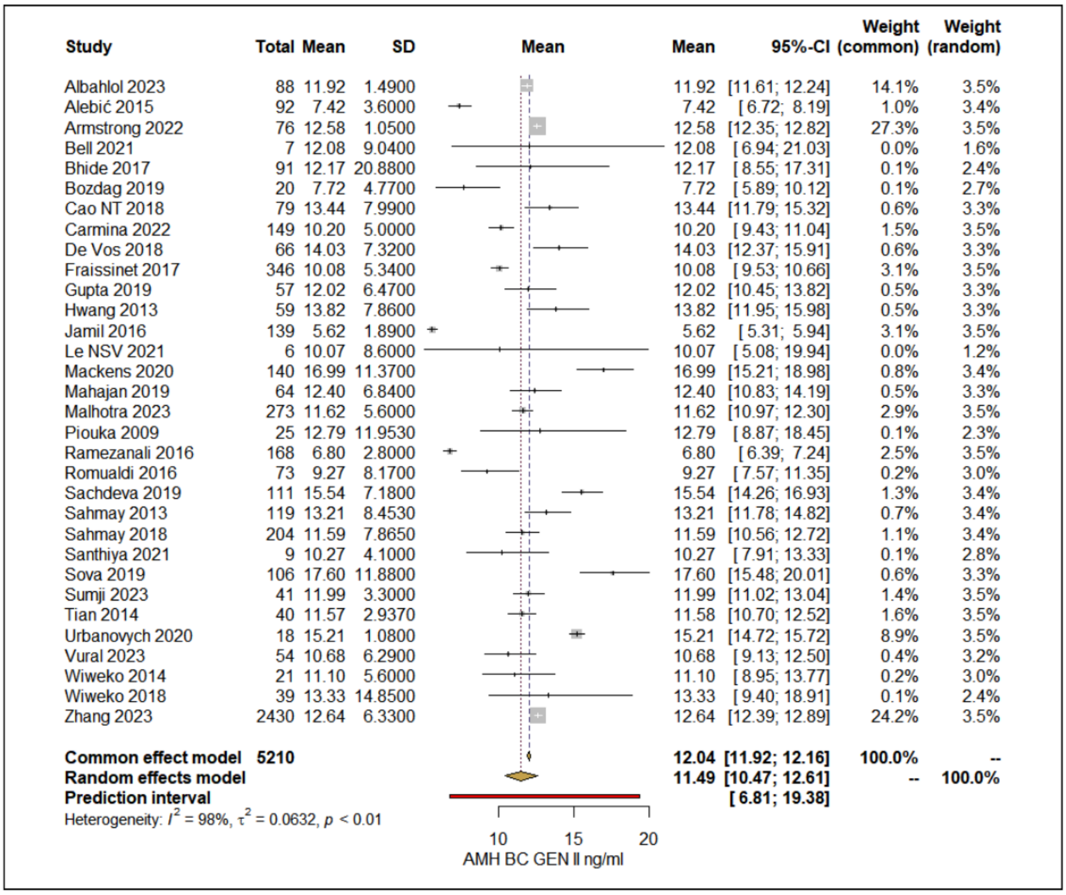
Study Total Mean SD Mean Mean 95%-CI Weight (common) Weight (random)
Albahlol 2023 88 11.92 1.4900 11.92 [11.61; 12.24] 14.1% 3.5%
Alebić 2015 92 7.42 3.6000 7.42 [6.72; 8.19] 1.0% 3.4%
Armstrong 2022 76 12.58 1.0500 12.58 [12.35; 12.82] 27.3% 3.5%
Bell 2021 7 12.08 9.0400 12.08 [6.94; 21.03] 0.0% 1.6%
Bhide 2017 91 12.17 20.8800 12.17 [8.55; 17.31] 0.1% 2.4%
Bozdag 2019 20 7.72 4.7700 7.72 [5.89; 10.12] 0.1% 2.7%
Cao NT 2018 79 13.44 7.9900 13.44 [11.79; 15.32] 0.6% 3.3%
Carmina 2022 149 10.20 5.0000 10.20 [9.43; 11.04] 1.5% 3.5%
De Vos 2018 66 14.03 7.3200 14.03 [12.37; 15.91] 0.6% 3.3%
Fraissinet 2017 346 10.08 5.3400 10.08 [9.53; 10.66] 3.1% 3.5%
Gupta 2019 57 12.02 6.4700 12.02 [10.45; 13.82] 0.5% 3.3%
Hwang 2013 59 13.82 7.8600 13.82 [11.95; 15.98] 0.5% 3.3%
Jamil 2016 139 5.62 1.8900 5.62 [5.31; 5.94] 3.1% 3.5%
Le NSV 2021 6 10.07 8.6000 10.07 [5.08; 19.94] 0.0% 1.2%
Mackens 2020 140 16.99 11.3700 16.99 [15.21; 18.98] 0.8% 3.4%
Mahajan 2019 64 12.40 6.8400 12.40 [10.83; 14.19] 0.5% 3.3%
Malhotra 2023 273 11.62 5.6000 11.62 [10.97; 12.30] 2.9% 3.5%
Piouka 2009 25 12.79 11.9530 12.79 [8.87; 18.45] 0.1% 2.3%
Ramezanali 2016 168 6.80 2.8000 6.80 [6.39; 7.24] 2.5% 3.5%
Romualdi 2016 73 9.27 8.1700 9.27 [7.57; 11.35] 0.2% 3.0%
Sachdeva 2019 111 15.54 7.1800 15.54 [14.26; 16.93] 1.3% 3.4%
Sahmay 2013 119 13.21 8.4530 13.21 [11.78; 14.82] 0.7% 3.4%
Sahmay 2018 204 11.59 7.8650 11.59 [10.56; 12.72] 1.1% 3.4%
Santhiya 2021 9 10.27 4.1000 10.27 [7.91; 13.33] 0.1% 2.8%
Sova 2019 106 17.60 11.8800 17.60 [15.48; 20.01] 0.6% 3.3%
Sumji 2023 41 11.99 3.3000 11.99 [11.02; 13.04] 1.4% 3.5%
Tian 2014 40 11.57 2.9370 11.58 [10.70; 12.52] 1.6% 3.5%
Urbanovych 2020 18 15.21 1.0800 15.21 [14.72; 15.72] 8.9% 3.5%
Vural 2023 54 10.68 6.2900 10.68 [9.13; 12.50] 0.4% 3.2%
Wiweko 2014 21 11.10 5.6000 11.10 [8.95; 13.77] 0.2% 3.0%
Wiweko 2018 39 13.33 14.8500 13.33 [9.40; 18.91] 0.1% 2.4%
Zhang 2023 2430 12.64 6.3300 12.64 [12.39; 12.89] 24.2% 3.5%
Common effect model 5210 12.04 [11.92; 12.16] 100.0% --
Random effects model 11.49 [10.47; 12.61] -- 100.0%
Prediction interval [6.81; 19.38]
Heterogeneity: I² = 98%, τ² = 0.0632, p < 0.01
10 15 20
AMH BC GEN II ng/ml

**PCOS-B Age:**

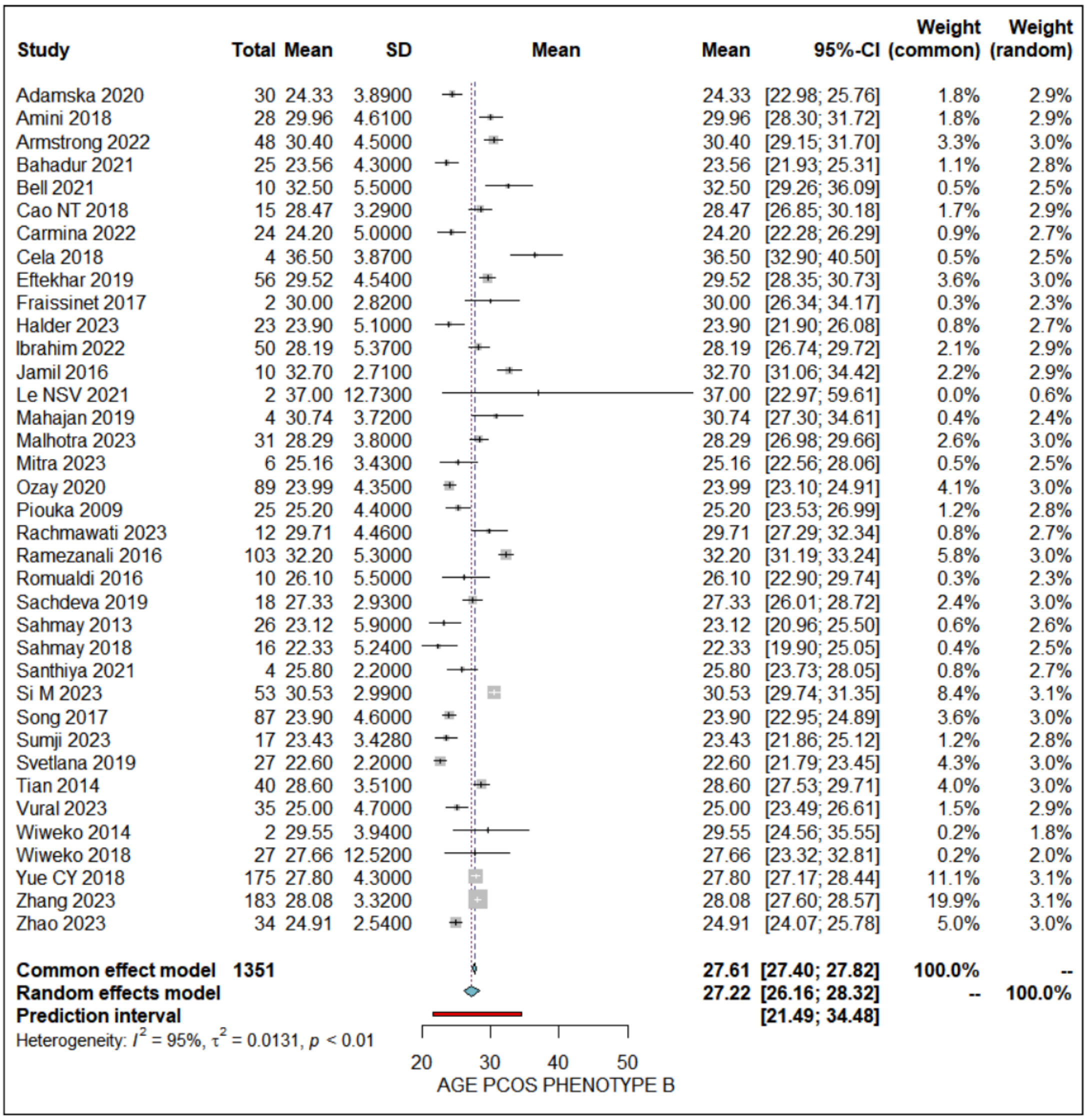

Study Total Mean SD Mean Mean 95%-CI Weight (common) Weight (random)
Adamska 2020 30 24.33 3.8900 24.33 [22.98; 25.76] 1.8% 2.9%
Amini 2018 28 29.96 4.6100 29.96 [28.30; 31.72] 1.8% 2.9%
Armstrong 2022 48 30.40 4.5000 30.40 [29.15; 31.70] 3.3% 3.0%
Bahadur 2021 25 23.56 4.3000 23.56 [21.93; 25.31] 1.1% 2.8%
Bell 2021 10 32.50 5.5000 32.50 [29.26; 36.09] 0.5% 2.5%
Cao NT 2018 15 28.47 3.2900 28.47 [26.85; 30.18] 1.7% 2.9%
Carmina 2022 24 24.20 5.0000 24.20 [22.28; 26.29] 0.9% 2.7%
Cela 2018 4 36.50 3.8700 36.50 [32.90; 40.50] 0.5% 2.5%
Eftekhar 2019 56 29.52 4.5400 29.52 [28.35; 30.73] 3.6% 3.0%
Fraissinet 2017 2 30.00 2.8200 30.00 [26.34; 34.17] 0.3% 2.3%
Halder 2023 23 23.90 5.1000 23.90 [21.90; 26.08] 0.8% 2.7%
Ibrahim 2022 50 28.19 5.3700 28.19 [26.74; 29.72] 2.1% 2.9%
Jamil 2016 10 32.70 2.7100 32.70 [31.06; 34.42] 2.2% 2.9%
Le NSV 2021 2 37.00 12.7300 37.00 [22.97; 59.61] 0.0% 0.6%
Mahajan 2019 4 30.74 3.7200 30.74 [27.30; 34.61] 0.4% 2.4%
Malhotra 2023 31 28.29 3.8000 28.29 [26.98; 29.66] 2.6% 3.0%
Mitra 2023 6 25.16 3.4300 25.16 [22.56; 28.06] 0.5% 2.5%
Ozay 2020 89 23.99 4.3500 23.99 [23.10; 24.91] 4.1% 3.0%
Piouka 2009 25 25.20 4.4000 25.20 [23.53; 26.99] 1.2% 2.8%
Rachmawati 2023 12 29.71 4.4600 29.71 [27.29; 32.34] 0.8% 2.7%
Ramezanali 2016 103 32.20 5.3000 32.20 [31.19; 33.24] 5.8% 3.0%
Romualdi 2016 10 26.10 5.5000 26.10 [22.90; 29.74] 0.3% 2.3%
Sachdeva 2019 18 27.33 2.9300 27.33 [26.01; 28.72] 2.4% 3.0%
Sahmay 2013 26 23.12 5.9000 23.12 [20.96; 25.50] 0.6% 2.6%
Sahmay 2018 16 22.33 5.2400 22.33 [19.90; 25.05] 0.4% 2.5%
Santhiya 2021 4 25.80 2.2000 25.80 [23.73; 28.05] 0.8% 2.7%
Si M 2023 53 30.53 2.9900 30.53 [29.74; 31.35] 8.4% 3.1%
Song 2017 87 23.90 4.6000 23.90 [22.95; 24.89] 3.6% 3.0%
Sumji 2023 17 23.43 3.4280 23.43 [21.86; 25.12] 1.2% 2.8%
Svetlana 2019 27 22.60 2.2000 22.60 [21.79; 23.45] 4.3% 3.0%
Tian 2014 40 28.60 3.5100 28.60 [27.53; 29.71] 4.0% 3.0%
Vural 2023 35 25.00 4.7000 25.00 [23.49; 26.61] 1.5% 2.9%
Wiweko 2014 2 29.55 3.9400 29.55 [24.56; 35.55] 0.2% 1.8%
Wiweko 2018 27 27.66 12.5200 27.66 [23.32; 32.81] 0.2% 2.0%
Yue CY 2018 175 27.80 4.3000 27.80 [27.17; 28.44] 11.1% 3.1%
Zhang 2023 183 28.08 3.3200 28.08 [27.60; 28.57] 19.9% 3.1%
Zhao 2023 34 24.91 2.5400 24.91 [24.07; 25.78] 5.0% 3.0%
Common effect model 1351 27.61 [27.40; 27.82] 100.0% --
Random effects model 27.22 [26.16; 28.32] -- 100.0%
Prediction interval [21.49; 34.48]
Heterogeneity: $I^2$ = 95%, $\tau^2$ = 0.0131, $p$ < 0.01
20 30 40 50
AGE PCOS PHENOTYPE B


**PCOS-B BMI:**

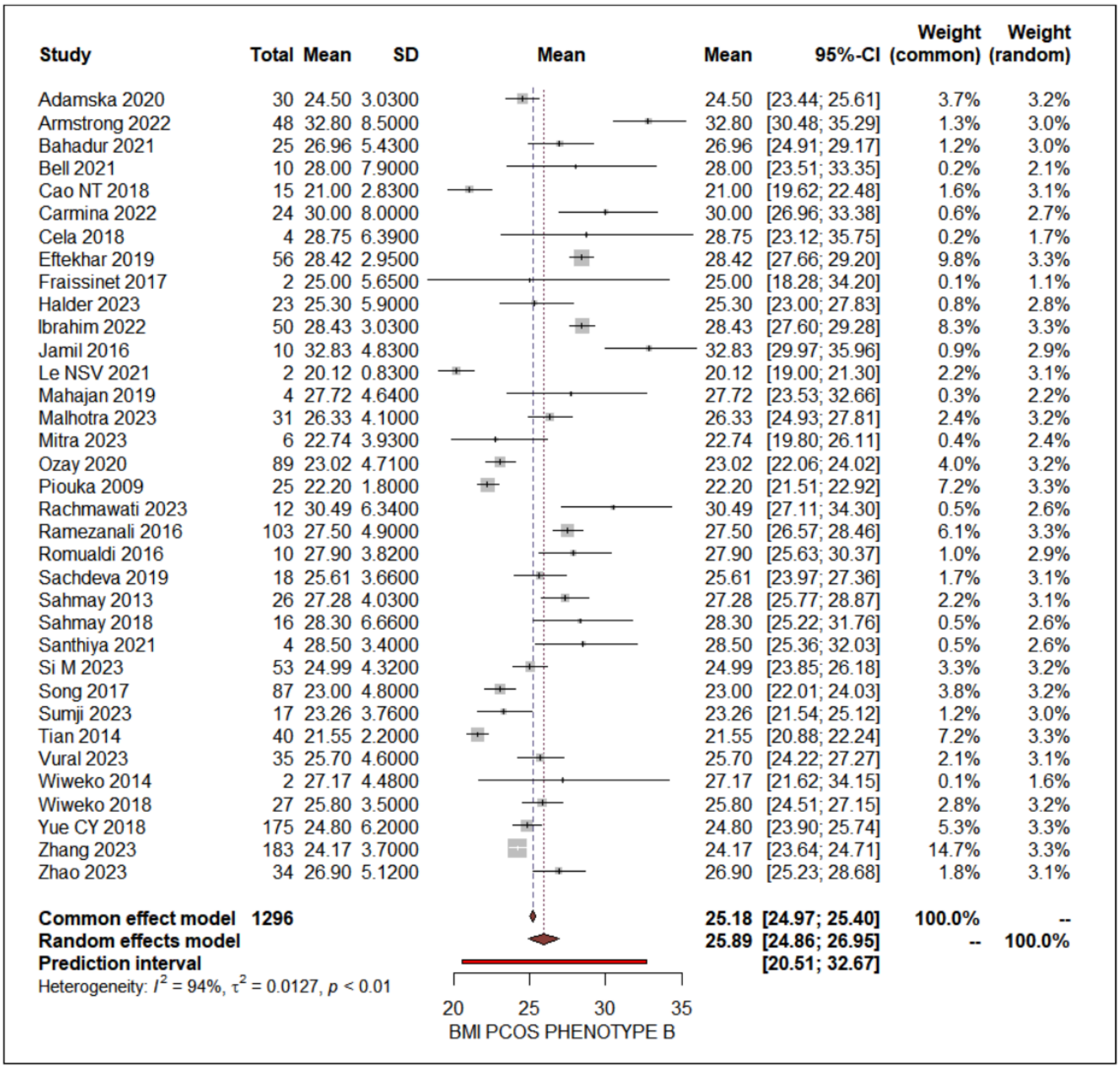


**PCOS-B AMH levels without assay differentiation:**

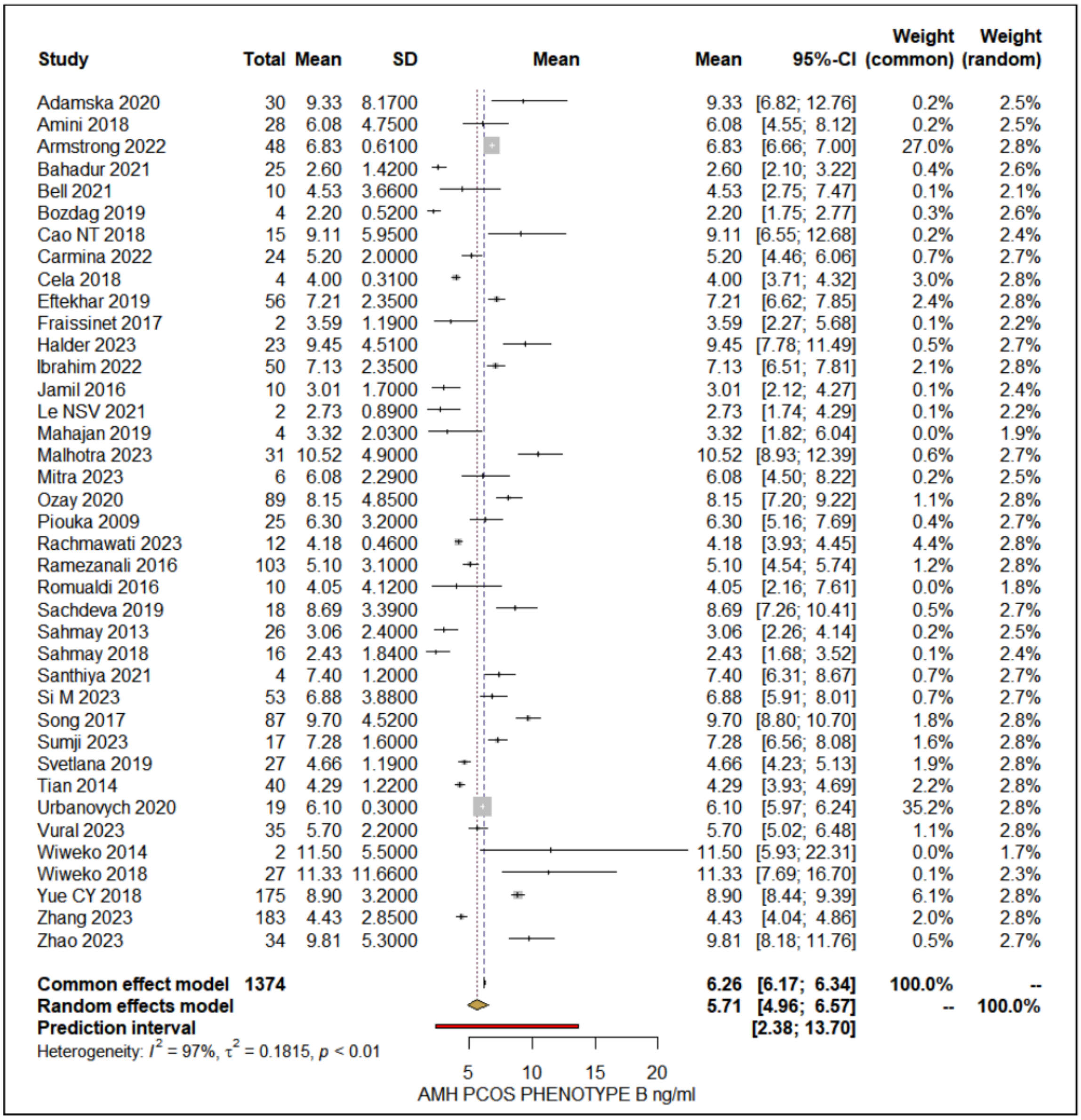


**PCOS-B AMH levels standardized to the Beckman Coulter Gen II assay:**

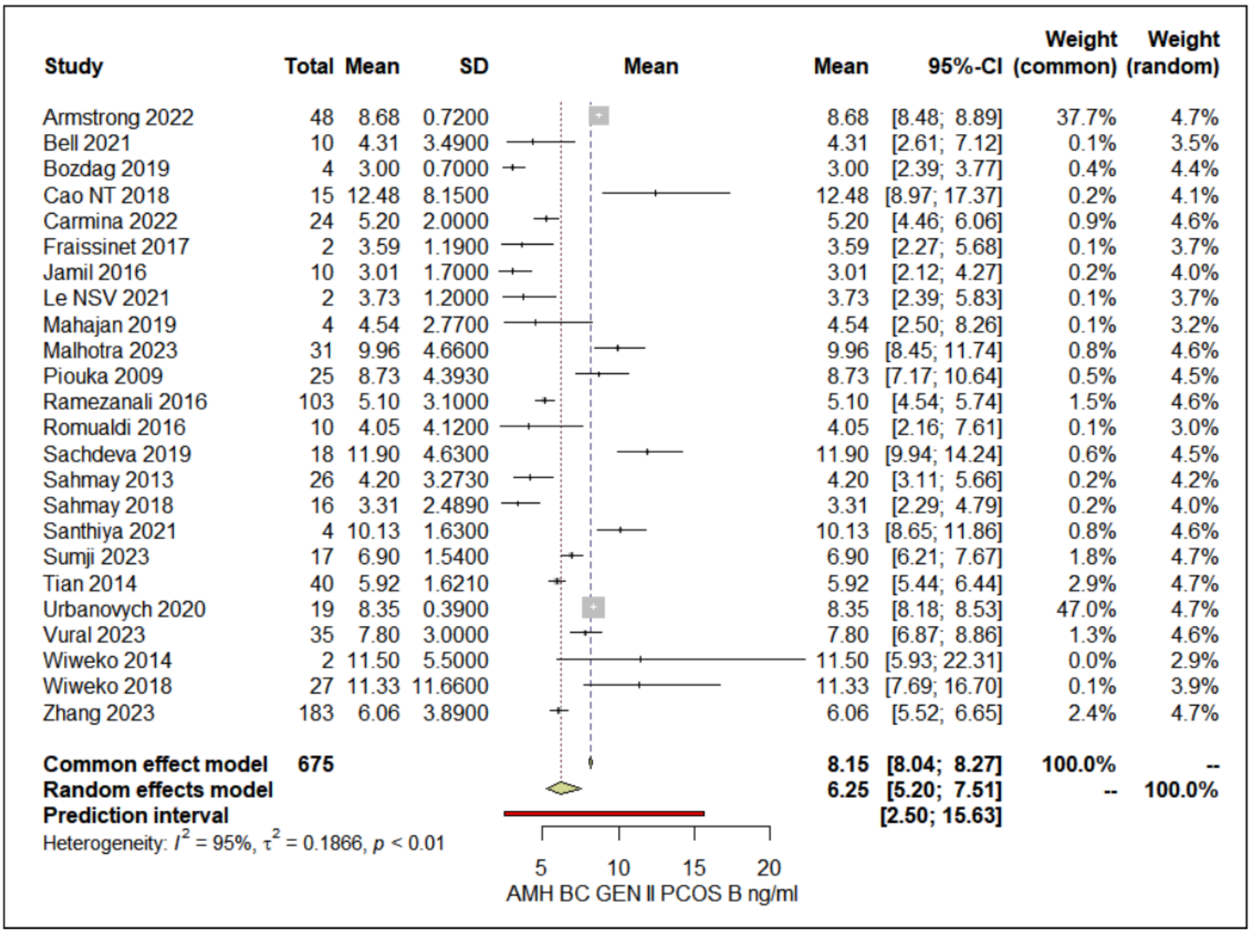
Study Total Mean SD Mean Mean 95%-CI Weight (common) Weight (random)
Armstrong 2022 48 8.68 0.7200 8.68 [8.48; 8.89] 37.7% 4.7%
Bell 2021 10 4.31 3.4900 4.31 [2.61; 7.12] 0.1% 3.5%
Bozdag 2019 4 3.00 0.7000 3.00 [2.39; 3.77] 0.4% 4.4%
Cao NT 2018 15 12.48 8.1500 12.48 [8.97; 17.37] 0.2% 4.1%
Carmina 2022 24 5.20 2.0000 5.20 [4.46; 6.06] 0.9% 4.6%
Fraissinet 2017 2 3.59 1.1900 3.59 [2.27; 5.68] 0.1% 3.7%
Jamil 2016 10 3.01 1.7000 3.01 [2.12; 4.27] 0.2% 4.0%
Le NSV 2021 2 3.73 1.2000 3.73 [2.39; 5.83] 0.1% 3.7%
Mahajan 2019 4 4.54 2.7700 4.54 [2.50; 8.26] 0.1% 3.2%
Malhotra 2023 31 9.96 4.6600 9.96 [8.45; 11.74] 0.8% 4.6%
Piouka 2009 25 8.73 4.3930 8.73 [7.17; 10.64] 0.5% 4.5%
Ramezanali 2016 103 5.10 3.1000 5.10 [4.54; 5.74] 1.5% 4.6%
Romualdi 2016 10 4.05 4.1200 4.05 [2.16; 7.61] 0.1% 3.0%
Sachdeva 2019 18 11.90 4.6300 11.90 [9.94; 14.24] 0.6% 4.5%
Sahmay 2013 26 4.20 3.2730 4.20 [3.11; 5.66] 0.2% 4.2%
Sahmay 2018 16 3.31 2.4890 3.31 [2.29; 4.79] 0.2% 4.0%
Santhiya 2021 4 10.13 1.6300 10.13 [8.65; 11.86] 0.8% 4.6%
Sumji 2023 17 6.90 1.5400 6.90 [6.21; 7.67] 1.8% 4.7%
Tian 2014 40 5.92 1.6210 5.92 [5.44; 6.44] 2.9% 4.7%
Urbanovych 2020 19 8.35 0.3900 8.35 [8.18; 8.53] 47.0% 4.7%
Vural 2023 35 7.80 3.0000 7.80 [6.87; 8.86] 1.3% 4.6%
Wiweko 2014 2 11.50 5.5000 11.50 [5.93; 22.31] 0.0% 2.9%
Wiweko 2018 27 11.33 11.6600 11.33 [7.69; 16.70] 0.1% 3.9%
Zhang 2023 183 6.06 3.8900 6.06 [5.52; 6.65] 2.4% 4.7%
Common effect model 675 8.15 [8.04; 8.27] 100.0% --
Random effects model 6.25 [5.20; 7.51] -- 100.0%
Prediction interval [2.50; 15.63]
Heterogeneity: $I^2$ = 95%, $\tau^2$ = 0.1866, $p$ < 0.01
5 10 15 20
AMH BC GEN II PCOS B ng/ml

**PCOS-C Age:**

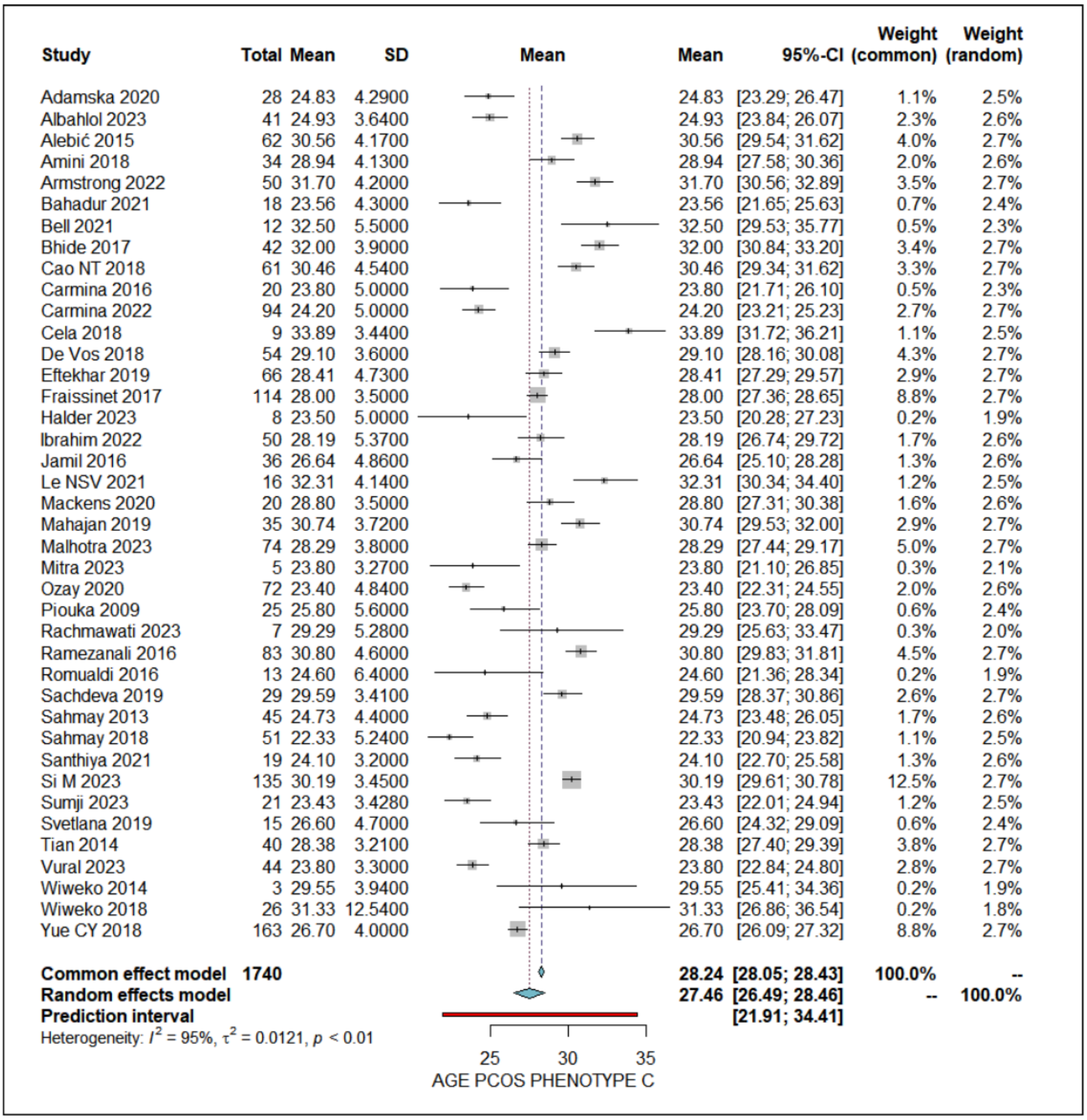
Study
Total Mean SD
Mean
Mean 95%-CI
Weight (common)
Weight (random)
Adamska 2020 28 24.83 4.2900 24.83 [23.29; 26.47] 1.1% 2.5%
Albahlol 2023 41 24.93 3.6400 24.93 [23.84; 26.07] 2.3% 2.6%
Alebić 2015 62 30.56 4.1700 30.56 [29.54; 31.62] 4.0% 2.7%
Amini 2018 34 28.94 4.1300 28.94 [27.58; 30.36] 2.0% 2.6%
Armstrong 2022 50 31.70 4.2000 31.70 [30.56; 32.89] 3.5% 2.7%
Bahadur 2021 18 23.56 4.3000 23.56 [21.65; 25.63] 0.7% 2.4%
Bell 2021 12 32.50 5.5000 32.50 [29.53; 35.77] 0.5% 2.3%
Bhide 2017 42 32.00 3.9000 32.00 [30.84; 33.20] 3.4% 2.7%
Cao NT 2018 61 30.46 4.5400 30.46 [29.34; 31.62] 3.3% 2.7%
Carmina 2016 20 23.80 5.0000 23.80 [21.71; 26.10] 0.5% 2.3%
Carmina 2022 94 24.20 5.0000 24.20 [23.21; 25.23] 2.7% 2.7%
Cela 2018 9 33.89 3.4400 33.89 [31.72; 36.21] 1.1% 2.5%
De Vos 2018 54 29.10 3.6000 29.10 [28.16; 30.08] 4.3% 2.7%
Eftekhar 2019 66 28.41 4.7300 28.41 [27.29; 29.57] 2.9% 2.7%
Fraissinet 2017 114 28.00 3.5000 28.00 [27.36; 28.65] 8.8% 2.7%
Halder 2023 8 23.50 5.0000 23.50 [20.28; 27.23] 0.2% 1.9%
Ibrahim 2022 50 28.19 5.3700 28.19 [26.74; 29.72] 1.7% 2.6%
Jamil 2016 36 26.64 4.8600 26.64 [25.10; 28.28] 1.3% 2.6%
Le NSV 2021 16 32.31 4.1400 32.31 [30.34; 34.40] 1.2% 2.5%
Mackens 2020 20 28.80 3.5000 28.80 [27.31; 30.38] 1.6% 2.6%
Mahajan 2019 35 30.74 3.7200 30.74 [29.53; 32.00] 2.9% 2.7%
Malhotra 2023 74 28.29 3.8000 28.29 [27.44; 29.17] 5.0% 2.7%
Mitra 2023 5 23.80 3.2700 23.80 [21.10; 26.85] 0.3% 2.1%
Ozay 2020 72 23.40 4.8400 23.40 [22.31; 24.55] 2.0% 2.6%
Piouka 2009 25 25.80 5.6000 25.80 [23.70; 28.09] 0.6% 2.4%
Rachmawati 2023 7 29.29 5.2800 29.29 [25.63; 33.47] 0.3% 2.0%
Ramezanali 2016 83 30.80 4.6000 30.80 [29.83; 31.81] 4.5% 2.7%
Romualdi 2016 13 24.60 6.4000 24.60 [21.36; 28.34] 0.2% 1.9%
Sachdeva 2019 29 29.59 3.4100 29.59 [28.37; 30.86] 2.6% 2.7%
Sahmay 2013 45 24.73 4.4000 24.73 [23.48; 26.05] 1.7% 2.6%
Sahmay 2018 51 22.33 5.2400 22.33 [20.94; 23.82] 1.1% 2.5%
Santhiya 2021 19 24.10 3.2000 24.10 [22.70; 25.58] 1.3% 2.6%
Si M 2023 135 30.19 3.4500 30.19 [29.61; 30.78] 12.5% 2.7%
Sumji 2023 21 23.43 3.4280 23.43 [22.01; 24.94] 1.2% 2.5%
Svetlana 2019 15 26.60 4.7000 26.60 [24.32; 29.09] 0.6% 2.4%
Tian 2014 40 28.38 3.2100 28.38 [27.40; 29.39] 3.8% 2.7%
Vural 2023 44 23.80 3.3000 23.80 [22.84; 24.80] 2.8% 2.7%
Wiweko 2014 3 29.55 3.9400 29.55 [25.41; 34.36] 0.2% 1.9%
Wiweko 2018 26 31.33 12.5400 31.33 [26.86; 36.54] 0.2% 1.8%
Yue CY 2018 163 26.70 4.0000 26.70 [26.09; 27.32] 8.8% 2.7%
Common effect model 1740 28.24 [28.05; 28.43] 100.0% --
Random effects model 27.46 [26.49; 28.46] -- 100.0%
Prediction interval [21.91; 34.41]
Heterogeneity: $I^2$ = 95%, $\tau^2$ = 0.0121, $p$ < 0.01
25 30 35
AGE PCOS PHENOTYPE C

**PCOS-C BMI:**

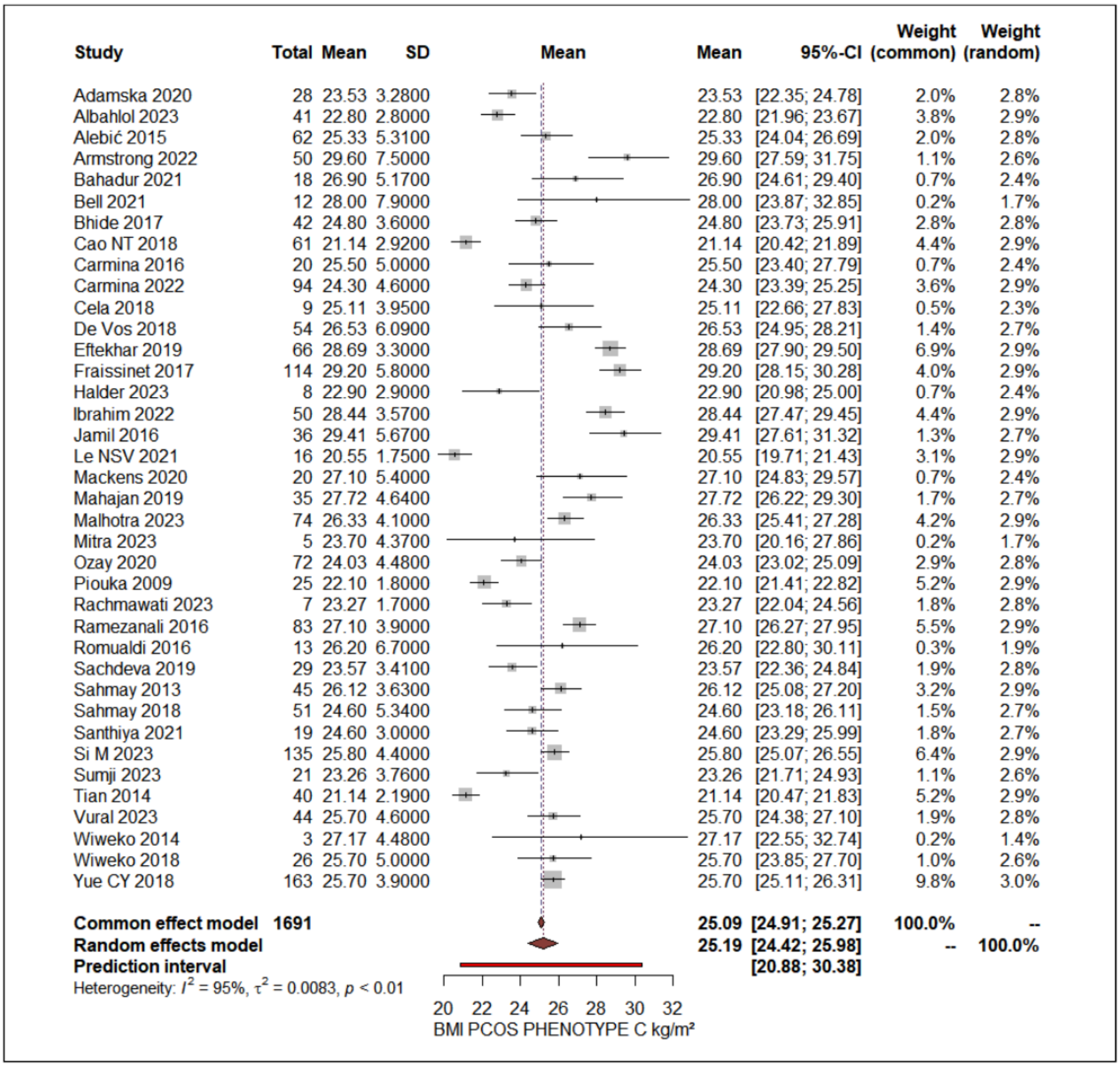


**PCOS-C AMH levels without assay differentiation:**

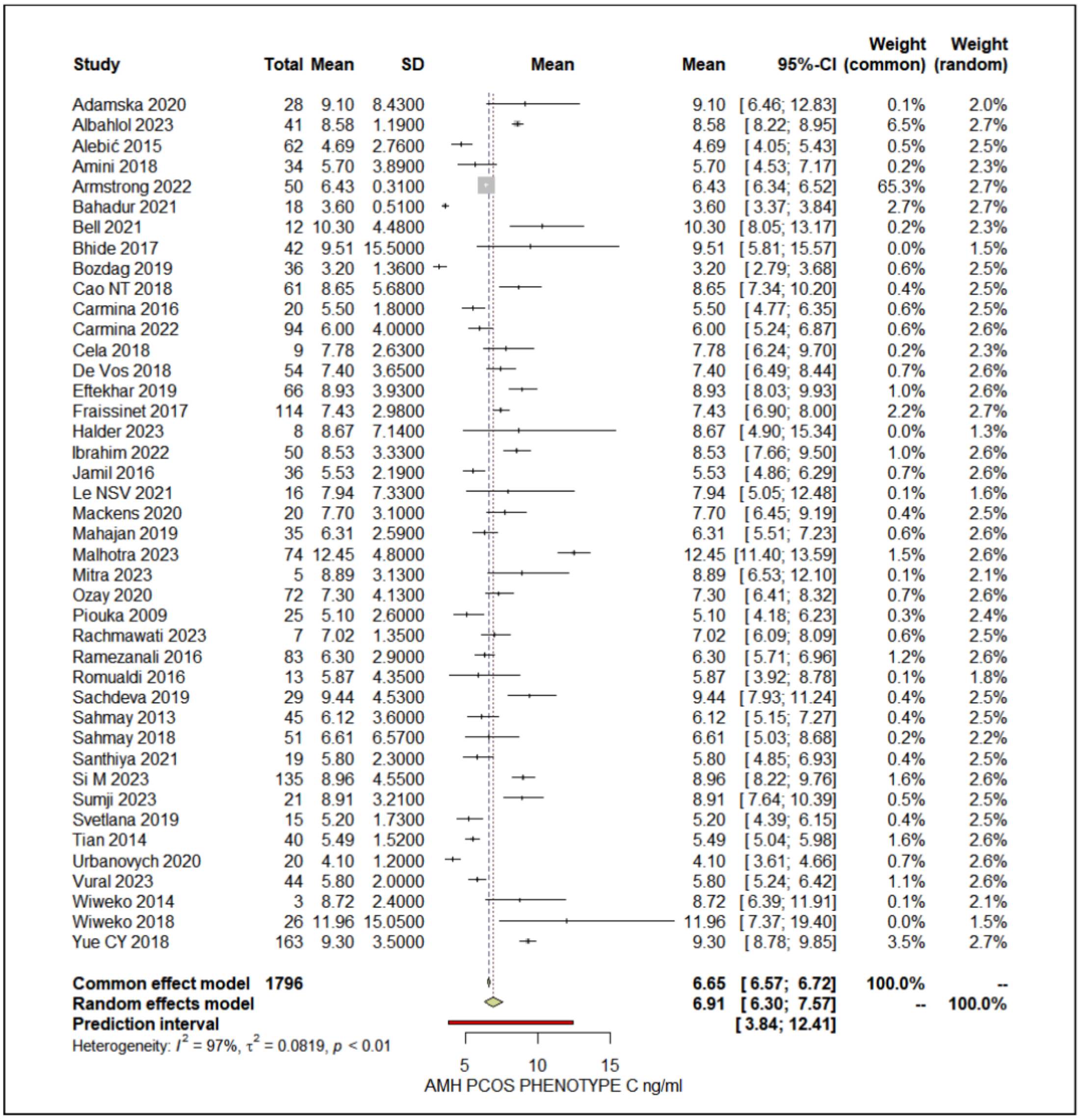


**PCOS-C AMH levels standardized to the Beckman Coulter Gen II assay:**

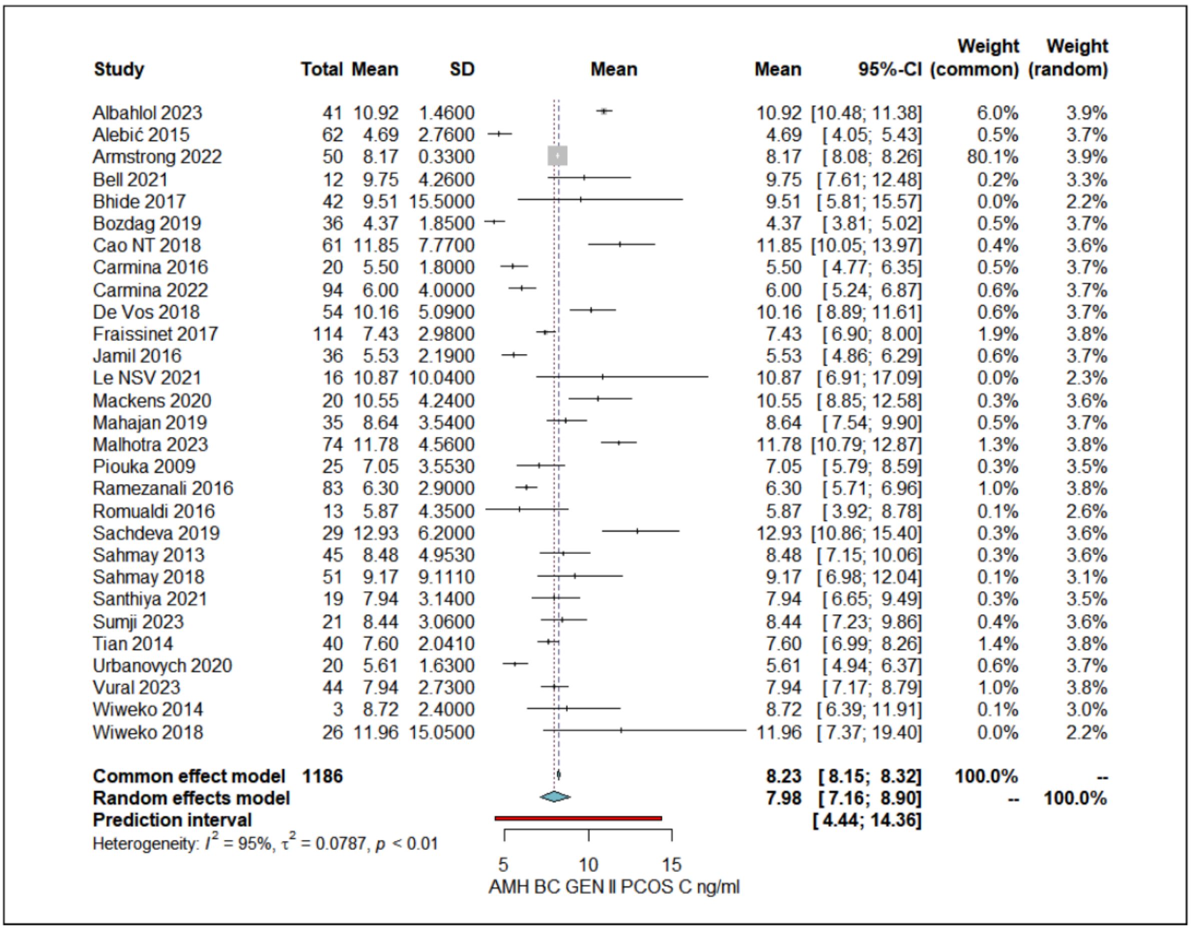
Study Total Mean SD Mean Mean 95%-CI Weight (common) Weight (random)
Albahlol 2023 41 10.92 1.4600 10.92 [10.48; 11.38] 6.0% 3.9%
Alebić 2015 62 4.69 2.7600 4.69 [ 4.05; 5.43] 0.5% 3.7%
Armstrong 2022 50 8.17 0.3300 8.17 [ 8.08; 8.26] 80.1% 3.9%
Bell 2021 12 9.75 4.2600 9.75 [ 7.61; 12.48] 0.2% 3.3%
Bhide 2017 42 9.51 15.5000 9.51 [ 5.81; 15.57] 0.0% 2.2%
Bozdag 2019 36 4.37 1.8500 4.37 [ 3.81; 5.02] 0.5% 3.7%
Cao NT 2018 61 11.85 7.7700 11.85 [10.05; 13.97] 0.4% 3.6%
Carmina 2016 20 5.50 1.8000 5.50 [ 4.77; 6.35] 0.5% 3.7%
Carmina 2022 94 6.00 4.0000 6.00 [ 5.24; 6.87] 0.6% 3.7%
De Vos 2018 54 10.16 5.0900 10.16 [ 8.89; 11.61] 0.6% 3.7%
Fraissinet 2017 114 7.43 2.9800 7.43 [ 6.90; 8.00] 1.9% 3.8%
Jamil 2016 36 5.53 2.1900 5.53 [ 4.86; 6.29] 0.6% 3.7%
Le NSV 2021 16 10.87 10.0400 10.87 [ 6.91; 17.09] 0.0% 2.3%
Mackens 2020 20 10.55 4.2400 10.55 [ 8.85; 12.58] 0.3% 3.6%
Mahajan 2019 35 8.64 3.5400 8.64 [ 7.54; 9.90] 0.5% 3.7%
Malhotra 2023 74 11.78 4.5600 11.78 [10.79; 12.87] 1.3% 3.8%
Piouka 2009 25 7.05 3.5530 7.05 [ 5.79; 8.59] 0.3% 3.5%
Ramezanali 2016 83 6.30 2.9000 6.30 [ 5.71; 6.96] 1.0% 3.8%
Romualdi 2016 13 5.87 4.3500 5.87 [ 3.92; 8.78] 0.1% 2.6%
Sachdeva 2019 29 12.93 6.2000 12.93 [10.86; 15.40] 0.3% 3.6%
Sahmay 2013 45 8.48 4.9530 8.48 [ 7.15; 10.06] 0.3% 3.6%
Sahmay 2018 51 9.17 9.1110 9.17 [ 6.98; 12.04] 0.1% 3.1%
Santhiya 2021 19 7.94 3.1400 7.94 [ 6.65; 9.49] 0.3% 3.5%
Sumji 2023 21 8.44 3.0600 8.44 [ 7.23; 9.86] 0.4% 3.6%
Tian 2014 40 7.60 2.0410 7.60 [ 6.99; 8.26] 1.4% 3.8%
Urbanovych 2020 20 5.61 1.6300 5.61 [ 4.94; 6.37] 0.6% 3.7%
Vural 2023 44 7.94 2.7300 7.94 [ 7.17; 8.79] 1.0% 3.8%
Wiweko 2014 3 8.72 2.4000 8.72 [ 6.39; 11.91] 0.1% 3.0%
Wiweko 2018 26 11.96 15.0500 11.96 [ 7.37; 19.40] 0.0% 2.2%
Common effect model 1186 8.23 [ 8.15; 8.32] 100.0% --
Random effects model 7.98 [ 7.16; 8.90] -- 100.0%
Prediction interval [ 4.44; 14.36]
Heterogeneity: $I^2$ = 95%, $\tau^2$ = 0.0787, $p$ < 0.01
5
10
15
AMH BC GEN II PCOS C ng/ml

**PCOS-D Age:**

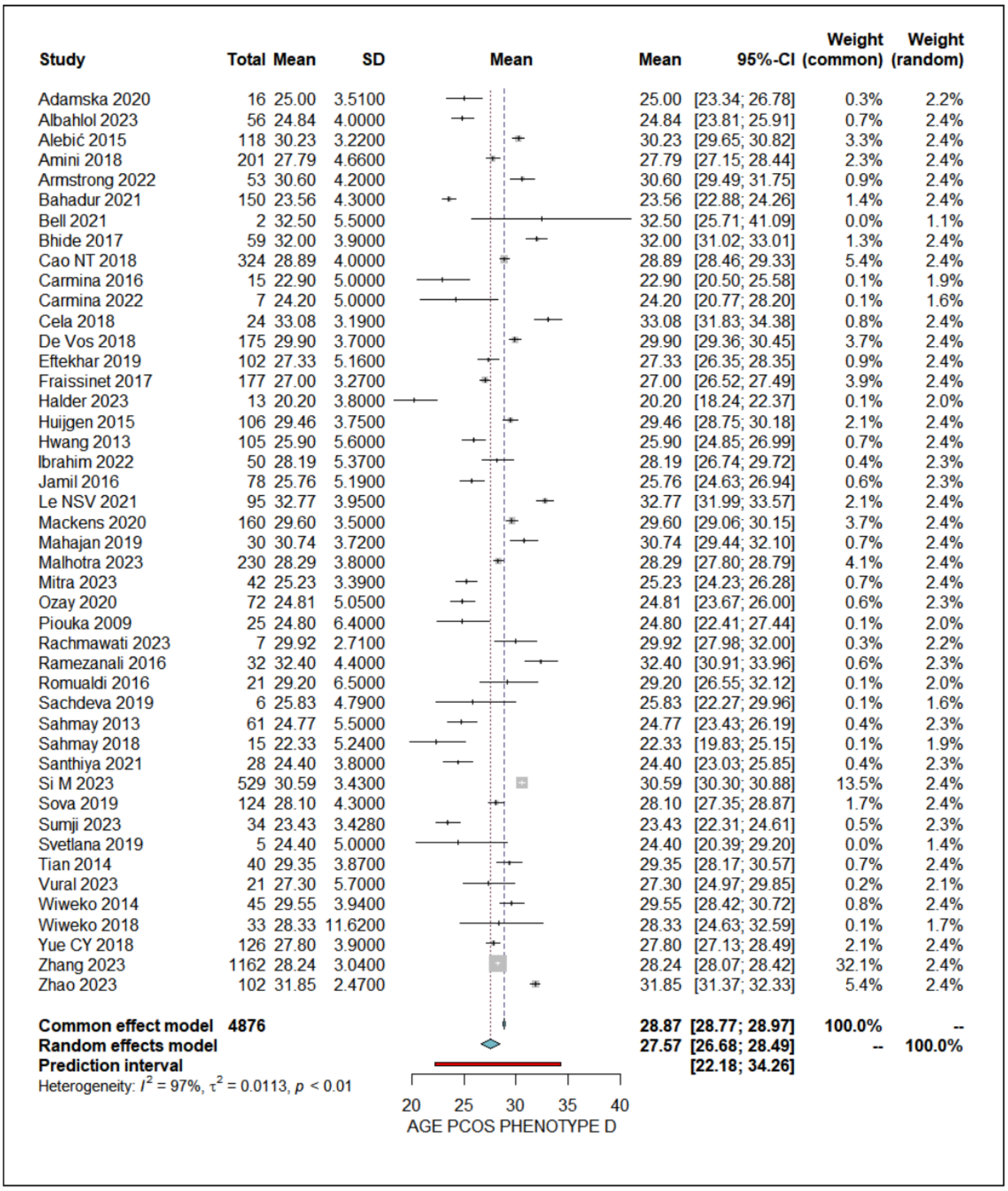
Study Total Mean SD Mean Mean 95%-CI Weight (common) Weight (random)
Adamska 2020 16 25.00 3.5100 25.00 [23.34; 26.78] 0.3% 2.2%
Albahlol 2023 56 24.84 4.0000 24.84 [23.81; 25.91] 0.7% 2.4%
Alebić 2015 118 30.23 3.2200 30.23 [29.65; 30.82] 3.3% 2.4%
Amini 2018 201 27.79 4.6600 27.79 [27.15; 28.44] 2.3% 2.4%
Armstrong 2022 53 30.60 4.2000 30.60 [29.49; 31.75] 0.9% 2.4%
Bahadur 2021 150 23.56 4.3000 23.56 [22.88; 24.26] 1.4% 2.4%
Bell 2021 2 32.50 5.5000 32.50 [25.71; 41.09] 0.0% 1.1%
Bhide 2017 59 32.00 3.9000 32.00 [31.02; 33.01] 1.3% 2.4%
Cao NT 2018 324 28.89 4.0000 28.89 [28.46; 29.33] 5.4% 2.4%
Carmina 2016 15 22.90 5.0000 22.90 [20.50; 25.58] 0.1% 1.9%
Carmina 2022 7 24.20 5.0000 24.20 [20.77; 28.20] 0.1% 1.6%
Cela 2018 24 33.08 3.1900 33.08 [31.83; 34.38] 0.8% 2.4%
De Vos 2018 175 29.90 3.7000 29.90 [29.36; 30.45] 3.7% 2.4%
Eftekhar 2019 102 27.33 5.1600 27.33 [26.35; 28.35] 0.9% 2.4%
Fraissinet 2017 177 27.00 3.2700 27.00 [26.52; 27.49] 3.9% 2.4%
Halder 2023 13 20.20 3.8000 20.20 [18.24; 22.37] 0.1% 2.0%
Huijgen 2015 106 29.46 3.7500 29.46 [28.75; 30.18] 2.1% 2.4%
Hwang 2013 105 25.90 5.6000 25.90 [24.85; 26.99] 0.7% 2.4%
Ibrahim 2022 50 28.19 5.3700 28.19 [26.74; 29.72] 0.4% 2.3%
Jamil 2016 78 25.76 5.1900 25.76 [24.63; 26.94] 0.6% 2.3%
Le NSV 2021 95 32.77 3.9500 32.77 [31.99; 33.57] 2.1% 2.4%
Mackens 2020 160 29.60 3.5000 29.60 [29.06; 30.15] 3.7% 2.4%
Mahajan 2019 30 30.74 3.7200 30.74 [29.44; 32.10] 0.7% 2.4%
Malhotra 2023 230 28.29 3.8000 28.29 [27.80; 28.79] 4.1% 2.4%
Mitra 2023 42 25.23 3.3900 25.23 [24.23; 26.28] 0.7% 2.4%
Ozay 2020 72 24.81 5.0500 24.81 [23.67; 26.00] 0.6% 2.3%
Piouka 2009 25 24.80 6.4000 24.80 [22.41; 27.44] 0.1% 2.0%
Rachmawati 2023 7 29.92 2.7100 29.92 [27.98; 32.00] 0.3% 2.2%
Ramezanali 2016 32 32.40 4.4000 32.40 [30.91; 33.96] 0.6% 2.3%
Romualdi 2016 21 29.20 6.5000 29.20 [26.55; 32.12] 0.1% 2.0%
Sachdeva 2019 6 25.83 4.7900 25.83 [22.27; 29.96] 0.1% 1.6%
Sahmay 2013 61 24.77 5.5000 24.77 [23.43; 26.19] 0.4% 2.3%
Sahmay 2018 15 22.33 5.2400 22.33 [19.83; 25.15] 0.1% 1.9%
Santhiya 2021 28 24.40 3.8000 24.40 [23.03; 25.85] 0.4% 2.3%
Si M 2023 529 30.59 3.4300 30.59 [30.30; 30.88] 13.5% 2.4%
Sova 2019 124 28.10 4.3000 28.10 [27.35; 28.87] 1.7% 2.4%
Sumji 2023 34 23.43 3.4280 23.43 [22.31; 24.61] 0.5% 2.3%
Svetlana 2019 5 24.40 5.0000 24.40 [20.39; 29.20] 0.0% 1.4%
Tian 2014 40 29.35 3.8700 29.35 [28.17; 30.57] 0.7% 2.4%
Vural 2023 21 27.30 5.7000 27.30 [24.97; 29.85] 0.2% 2.1%
Wiweko 2014 45 29.55 3.9400 29.55 [28.42; 30.72] 0.8% 2.4%
Wiweko 2018 33 28.33 11.6200 28.33 [24.63; 32.59] 0.1% 1.7%
Yue CY 2018 126 27.80 3.9000 27.80 [27.13; 28.49] 2.1% 2.4%
Zhang 2023 1162 28.24 3.0400 28.24 [28.07; 28.42] 32.1% 2.4%
Zhao 2023 102 31.85 2.4700 31.85 [31.37; 32.33] 5.4% 2.4%
Common effect model 4876 28.87 [28.77; 28.97] 100.0% --
Random effects model 27.57 [26.68; 28.49] -- 100.0%
Prediction interval [22.18; 34.26]
Heterogeneity: $I^2$ = 97%, $\tau^2$ = 0.0113, $p$ < 0.01
20 25 30 35 40
AGE PCOS PHENOTYPE D

**PCOS-D BMI:**

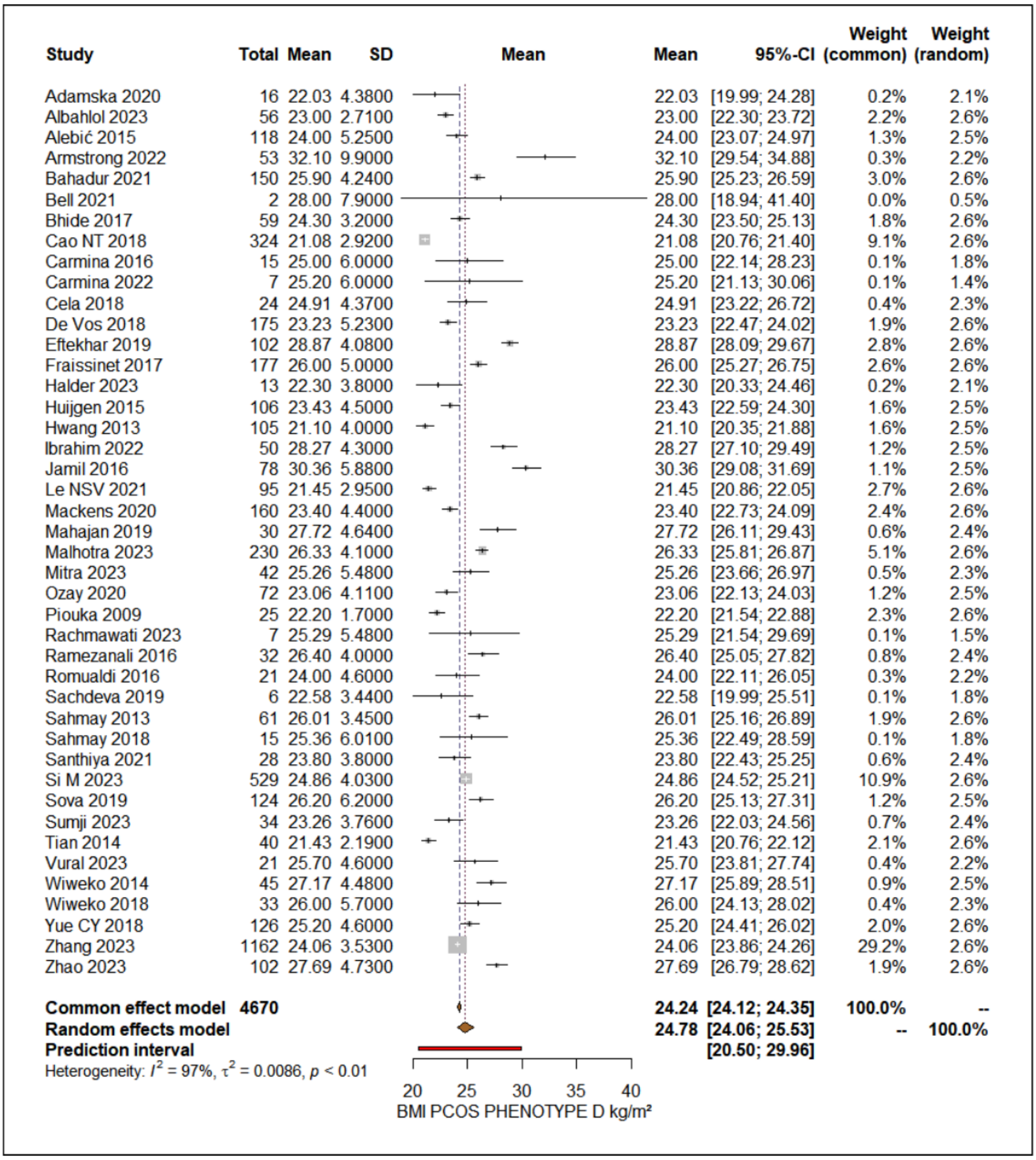


**PCOS-D AMH levels without assay differentiation:**

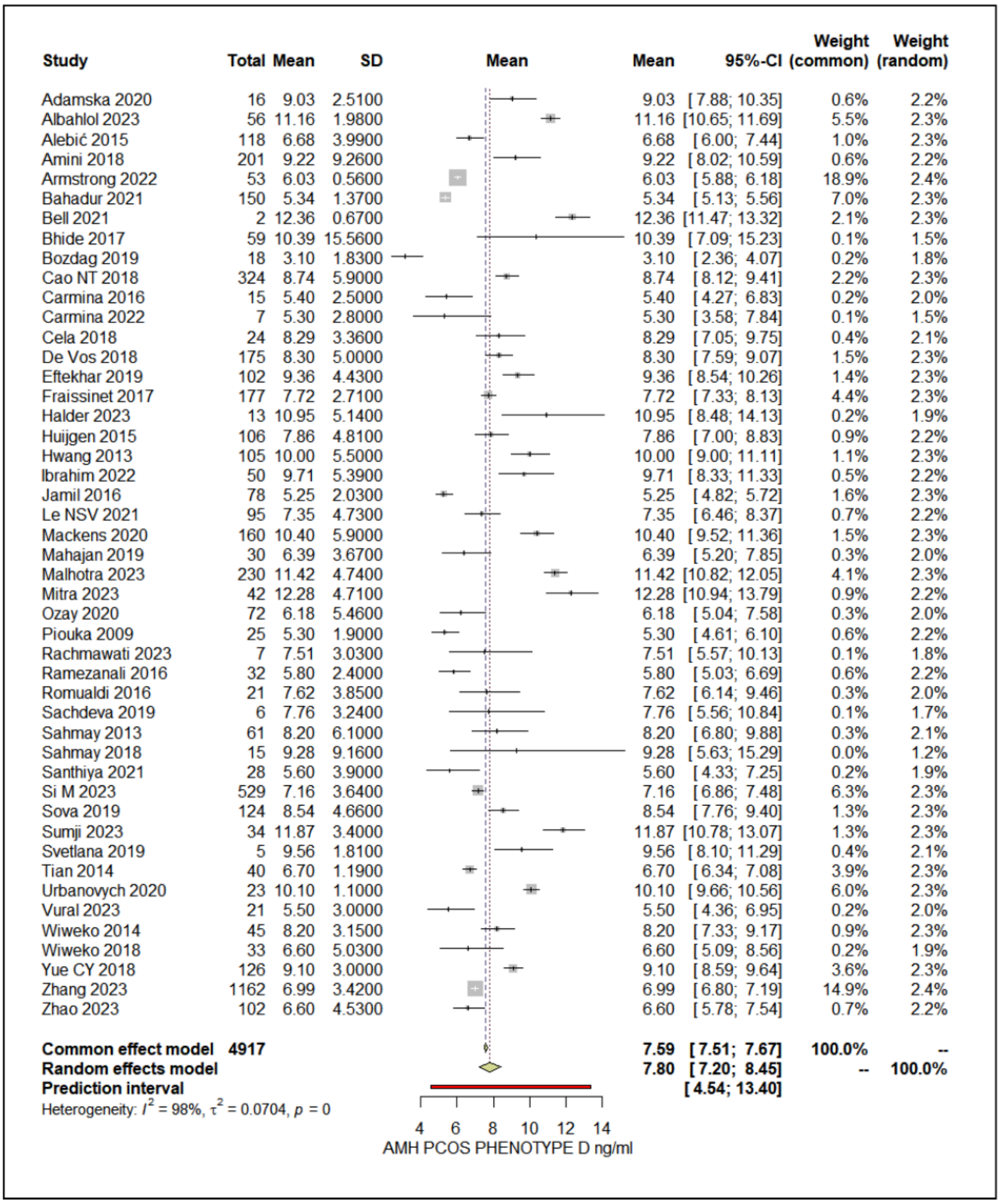


**PCOS-D AMH levels standardized to the Beckman Coulter Gen II assay:**

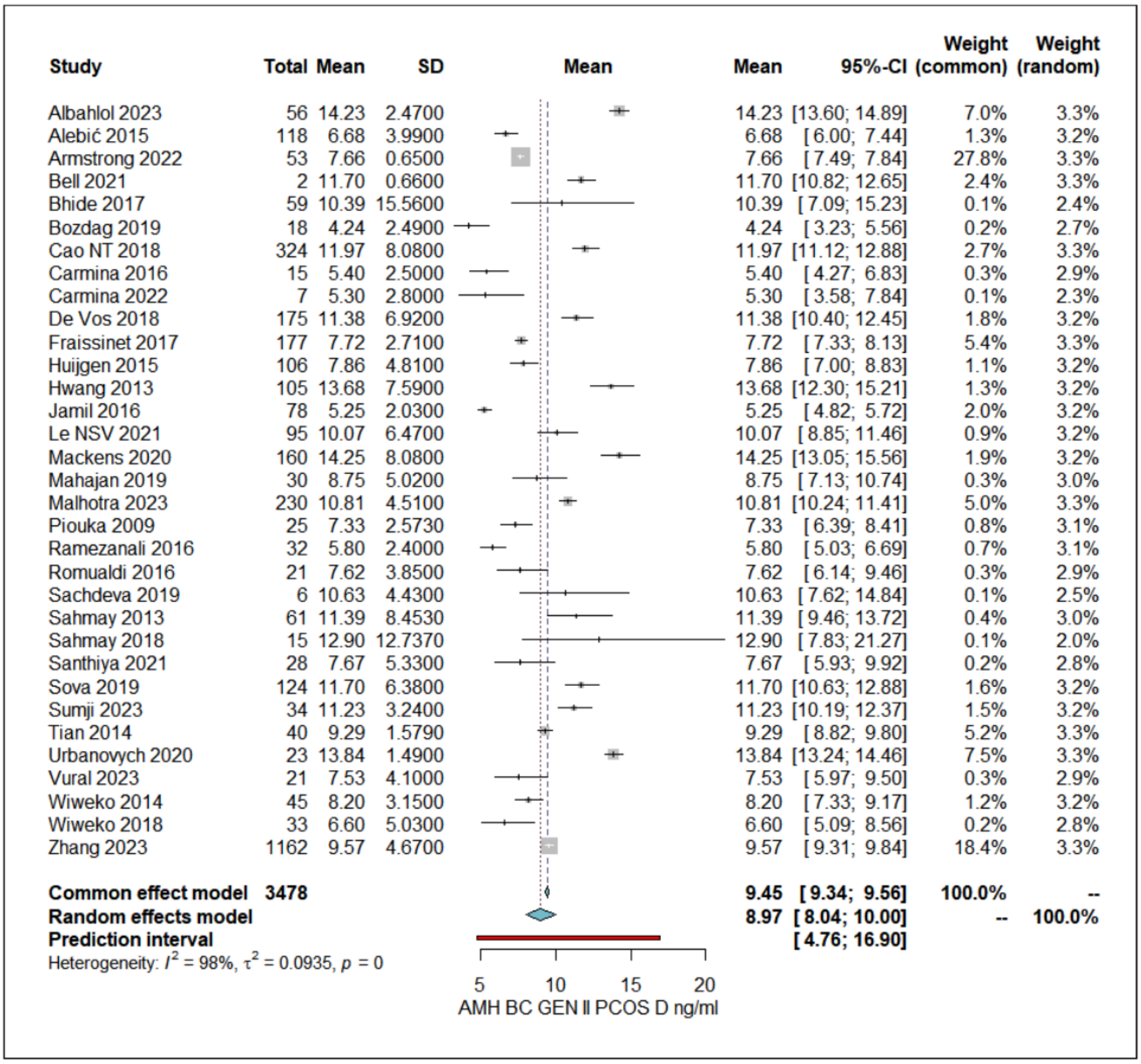
Study
Total
Mean
SD
Mean
Mean
95%-CI
Weight (common)
Weight (random)
Albahlol 2023 56 14.23 2.4700 14.23 [13.60; 14.89] 7.0% 3.3%
Alebić 2015 118 6.68 3.9900 6.68 [6.00; 7.44] 1.3% 3.2%
Armstrong 2022 53 7.66 0.6500 7.66 [7.49; 7.84] 27.8% 3.3%
Bell 2021 2 11.70 0.6600 11.70 [10.82; 12.65] 2.4% 3.3%
Bhide 2017 59 10.39 15.5600 10.39 [7.09; 15.23] 0.1% 2.4%
Bozdag 2019 18 4.24 2.4900 4.24 [3.23; 5.56] 0.2% 2.7%
Cao NT 2018 324 11.97 8.0800 11.97 [11.12; 12.88] 2.7% 3.3%
Carmina 2016 15 5.40 2.5000 5.40 [4.27; 6.83] 0.3% 2.9%
Carmina 2022 7 5.30 2.8000 5.30 [3.58; 7.84] 0.1% 2.3%
De Vos 2018 175 11.38 6.9200 11.38 [10.40; 12.45] 1.8% 3.2%
Fraissinet 2017 177 7.72 2.7100 7.72 [7.33; 8.13] 5.4% 3.3%
Huijgen 2015 106 7.86 4.8100 7.86 [7.00; 8.83] 1.1% 3.2%
Hwang 2013 105 13.68 7.5900 13.68 [12.30; 15.21] 1.3% 3.2%
Jamil 2016 78 5.25 2.0300 5.25 [4.82; 5.72] 2.0% 3.2%
Le NSV 2021 95 10.07 6.4700 10.07 [8.85; 11.46] 0.9% 3.2%
Mackens 2020 160 14.25 8.0800 14.25 [13.05; 15.56] 1.9% 3.2%
Mahajan 2019 30 8.75 5.0200 8.75 [7.13; 10.74] 0.3% 3.0%
Malhotra 2023 230 10.81 4.5100 10.81 [10.24; 11.41] 5.0% 3.3%
Piouka 2009 25 7.33 2.5730 7.33 [6.39; 8.41] 0.8% 3.1%
Ramezanali 2016 32 5.80 2.4000 5.80 [5.03; 6.69] 0.7% 3.1%
Romualdi 2016 21 7.62 3.8500 7.62 [6.14; 9.46] 0.3% 2.9%
Sachdeva 2019 6 10.63 4.4300 10.63 [7.62; 14.84] 0.1% 2.5%
Sahmay 2013 61 11.39 8.4530 11.39 [9.46; 13.72] 0.4% 3.0%
Sahmay 2018 15 12.90 12.7370 12.90 [7.83; 21.27] 0.1% 2.0%
Santhiya 2021 28 7.67 5.3300 7.67 [5.93; 9.92] 0.2% 2.8%
Sova 2019 124 11.70 6.3800 11.70 [10.63; 12.88] 1.6% 3.2%
Sumji 2023 34 11.23 3.2400 11.23 [10.19; 12.37] 1.5% 3.2%
Tian 2014 40 9.29 1.5790 9.29 [8.82; 9.80] 5.2% 3.3%
Urbanovych 2020 23 13.84 1.4900 13.84 [13.24; 14.46] 7.5% 3.3%
Vural 2023 21 7.53 4.1000 7.53 [5.97; 9.50] 0.3% 2.9%
Wiweko 2014 45 8.20 3.1500 8.20 [7.33; 9.17] 1.2% 3.2%
Wiweko 2018 33 6.60 5.0300 6.60 [5.09; 8.56] 0.2% 2.8%
Zhang 2023 1162 9.57 4.6700 9.57 [9.31; 9.84] 18.4% 3.3%
Common effect model 3478 9.45 [9.34; 9.56] 100.0% --
Random effects model 8.97 [8.04; 10.00] -- 100.0%
Prediction interval [4.76; 16.90]
Heterogeneity: $I^2$ = 98%, $\tau^2$ = 0.0935, $p$ = 0
5 10 15 20
AMH BC GEN II PCOS D ng/ml
